# SARS-CoV-2 Seroprevalence in Tamil Nadu in October-November 2020

**DOI:** 10.1101/2021.02.03.21250949

**Authors:** Anup Malani, Sabareesh Ramachandran, Vaidehi Tandel, Rajeswari Parasa, Sofia Imad, S. Sudharshini, V. Prakash, Y. Yogananth, S. Raju, T.S. Selvavinayagam

## Abstract

A population-representative serological study was conducted in all districts of the state of Tamil Nadu (population 72 million), India, in October-November 2020. State-level seroprevalence was 31.6%. However, this masks substantial variation across the state. Seroprevalence ranged from just 11.1% in The Nilgris to 51.0% in Perambalur district. Seroprevalence in urban areas (36.9%) was higher than in rural areas (26.9%). Females (30.8%) had similar seroprevalence to males (30.3%). However, working age populations (age 40-49: 31.6%) have significantly higher seroprevalence than the youth (age 18-29: 30.7%) or elderly (age 70+: 25.8%). Estimated seroprevalence implies that at least 22.6 million persons were infected by the end of November, roughly 36 times the number of confirmed cases. Estimated seroprevalence implies an infection fatality rate of 0.052%.

## Introduction

Knowledge of population-level immunity is critical for understanding the epidemiology of SARS-CoV-2 (COVID-19) and formulating effective infection control, including the allocation of scarce vaccines. Tamil Nadu is the 6th most populous state in India, with roughly 72 million persons^1^. It has reported roughly 820,000 COVID-19 cases and 12,000 deaths, ranked 4th and 2nd highest, respectively, among Indian states^2^. Reported cases are not, however, gathered from population-representative samples.

The state conducted a population-level seroprevalence survey of 26,640 adults across the 37 districts of the state in October-November 2020. We report seroprevalence estimates from this survey by district, by demographic groups, and by urban status. We also compare the results of the survey to estimates from reported cases to measure the degree to which serological surveys underestimate population immunity.

## Methods

This study was approved by the Directorate of Public Health & Preventative Medicine, Government of Tamil Nadu, and the Institutional Ethics Committee of Madras Medical College, Chennai, India.

### Outcomes

The first primary endpoint of the study is the rate of positive results on CLIA antibody tests at the district-level. The second primary endpoint is seroprevalence rates at the district-level once seropositivity rates are adjusted for test inaccuracy.

There are multiple secondary endpoints. One set is seroprevalence (a) by age categories and sex, (b) by rural or urban status, and (c) at the state level. A final secondary outcome is the difference between population immunity estimated by serological survey and by reported cases.

### Sample and location

Individuals residing in Tamil Nadu and ages 18 years and older were eligible for this study. The exclusion criteria were refusal to consent and contraindication to venipuncture.

### Sample size

The state sought to sample roughly 300 persons per 1 million population based on the 2011 Indian Census. The state’s population is organized into districts, districts are organized into health unit districts (HUD), and HUDs are organized into clusters, defined as a street in urban areas and habitations in rural areas. Within each cluster the study aimed to sample 30 individuals. Therefore, the sample rate can be converted into a target number of clusters per HUD. In total 888 clusters were sampled.

### Sample selection

The study selected participants within a HUD in three steps. First, within each HUD, the study randomly selected clusters. Second, within each cluster, we selected a random GPS starting point. Third, we sampled one participant from households adjacent to that starting point until 30 persons consented within a cluster. Within each household, the member asked to provide a biosample was selected via the Kish method^3^.

### Data collection and timing

Data was collected between October 19 - November 30, 2020. Each participant was asked to complete a health questionnaire and provide 5ml venous blood collected in EDTA vacutainers. Serum was analyzed for IgG antibodies to the SARS-CoV-2 spike protein using either the iFlash-SARS-CoV-2 IgG (Shenzhen YHLO Biotech; sensitivity of 95.9% and specificity of 95.7% per manufacturer)^4^ or the Vitros anti-SARS-CoV-2 IgG CLIA kit (Ortho-Clinical Diagnostics; sensitivity of 90% and specificity of 100% per manufacturer)^5^. Clusters were characterized as rural if they were in Census-defined villages. The government shared data on each reported COVID-19 case and death, including date and demographics.

### Statistical analysis

We estimate the proportion of positive CLIA tests by district by estimating a weighted logit regression of test result on district indicators and reporting the inverse logit of the coefficient for each district indicator. Observations are weighted by the inverse of sampling probability for their age and gender groups. Standard errors calculated account for correlations at the cluster level.

We estimate the seroprevalence by district in two steps. First, we calculate the weighted proportion of positive tests at the district level everywhere except Chennai, where we calculate it at the health unit district (HUD), a subset of districts. All samples in a district use the same CLIA kit, except in Chennai, where all samples in a HUD do so. We estimate a weighted logit regressions of test results on district indicators outside Chennai and HUD indicators in Chennai and take the inverse logit of the coefficient for each jurisdiction indicator. Observations are weighted by the inverse of sampling probability for their age and gender groups. Standard errors are clustered at the cluster level. Second, for each jurisdiction, we predict seroprevalence using the Rogan-Gladden formula, test parameters for the kit used in each jurisdiction, and regression estimates of seropositive proportion by jurisdiction. In Chennai district, we calculate seroprevalence at the district level as a weighted average of seroprevalence at the HUD level using as weights the share of clusters in each HUD. (We employ this approach to Chennai in the estimators below.)

We estimate the seroprevalence in the state by aggregating the seroprevalence across districts weighted by 2011 census data on the relative populations of districts.

We estimate seroprevalence by demographic group in three steps. First, we calculate the proportion of positive tests at the jurisdiction-by-demographic group level using logit regressions of test results on jurisdiction-by-demographic group indicators. Demographic groups indicators are sex x age for 6 age bins. Standard errors are clustered at the cluster level. Second, we predict district-by-demographic group level seroprevalence using the Rogan-Gladden formula. Third, we compute the weighted average of seroprevalence at the demographic-group level using as weights the share of demographic-group population in each district using data from the 2011 Indian census.

We estimate seroprevalence by urban status in the same manner we estimate it for demographic groups with two changes. First, we use the urban status of a cluster in lieu of demographic status of an individual at each step. Second, observations in our regression are weighted by inverse of the sampling probability for their urban status.

We estimate the infection fatality rate (IFR) for a population (defined by state, district, demographic group, or urban status) by dividing total SARS-CoV-2-related deaths reported as of two days after the last date of sampling in a district by the estimated size of previously infected persons in that population. We obtain data on deaths by district and demographic group from daily reports from the Tamil Nadu government. The date of the death count reflects that fact that the delay the delay between infection and death is on average two days longer than the delay between infection and seroconversion.^6, 7^

We estimate the size of a population that was previously infected by multiplying our seroprevalence estimates for that population by the size of those populations as reported in the 2011 census. We estimate the degree of undercounting of cases in a population by dividing the estimated number previously infected in the population by the number of officially reported cases in that population as of 1 week before the median sampling date (median October 23, 2020). We obtain data on officially reported cases from the government of Tamil Nadu. The lag accounts for the delay both between infection and seropositive status and between infection and prevalence testing.

We calculate the Pearson’s correlation coefficient between IFR and age at the individual level, between undercounting rate and testing rate (tests per million as of median date of testing) by district, and between the number of SARS-CoV-2-related deaths and the testing rate by district.

Statistical tests comparing groups are performed using a two-sided Wald test with 95%.

All statistical analyses were conducted with Microsoft Excel 365 (Microsoft, USA) and Stata 16 (StataCorp, USA). All plots were generated in R.

## Results

One person aged 16 was incorrectly consented and dropped from the analysis. The study was unable to obtain lab results for 287 additional observations. The study employs 26,135 test results. Because numerous clusters had fewer than 30 persons consenting, the study yielded 292 samples per million population.

Table 1 reports the demographic characteristics of the sample. Age is missing for 8 persons and 6 persons are transgender. The sample has substantially more females and fewer persons age 18-29 than the general population.

**Table 1.** Demographics of sample, as compared to 2011 Census.

|  |  | **Sample** |  |  | **2011**<br>**Census** |
| --- | --- | --- | --- | --- | --- |
|  |  | **Mean** | **Lower**<br>**bound** | **Upper**<br>**bound** |  |
| Gender | Female | 61% | 60% | 61% | 50% |
|  | Male | 39% | 39% | 40% | 50% |
| Age | 18-29 | 23% | 22% | 23% | 27% |
|  | 30-39 | 23% | 23% | 24% | 24% |
|  | 40-49 | 20% | 20% | 21% | 20% |
|  | 50-59 | 16% | 16% | 17% | 14% |
|  | 60-69 | 11% | 11% | 12% | 10% |
|  | 70+ | 6% | 6% | 6% | 6% |
| Obs. |  | 26,135 |  |  | 72,147,030 |

Note. Census 2011 number for ages 18-29 includes only those ages 20-29.

Seropositivity varies dramatically across the state, from 12.1% in The Nilgris to 49.3% in Perambalur district (Figure 1). Seroprevalence has a similar pattern to seropositivity (Figure 2). Seroprevalence is significantly greater in urban areas (36.9%; rural, 26.9%; p<0.001) (Table 2). State level seroprevalence is 31.6% (95% CI: 30.4-32.8%).

**Figure 1.**
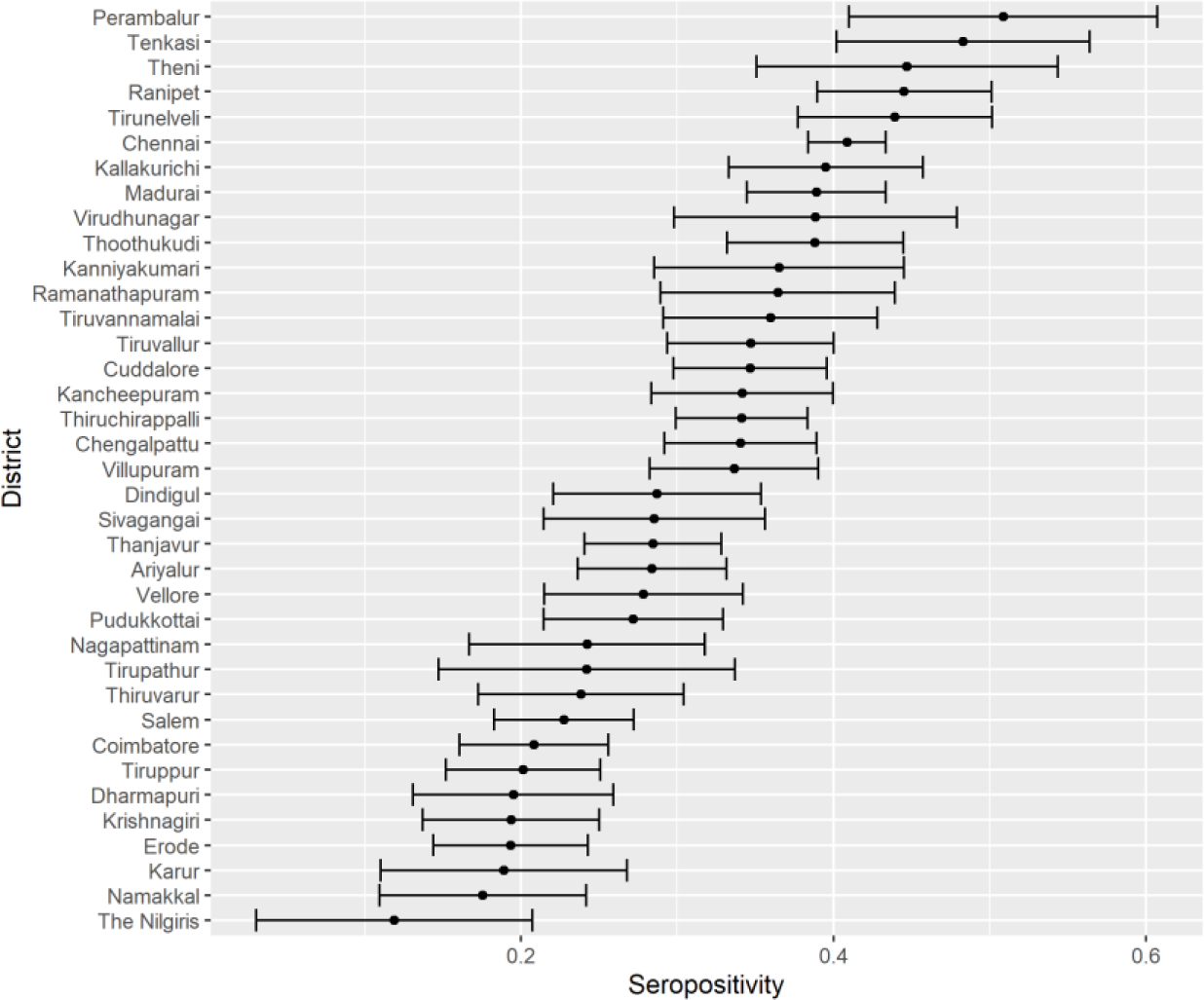
Proportion of positive CLIA tests by district.

**Figure 2.**
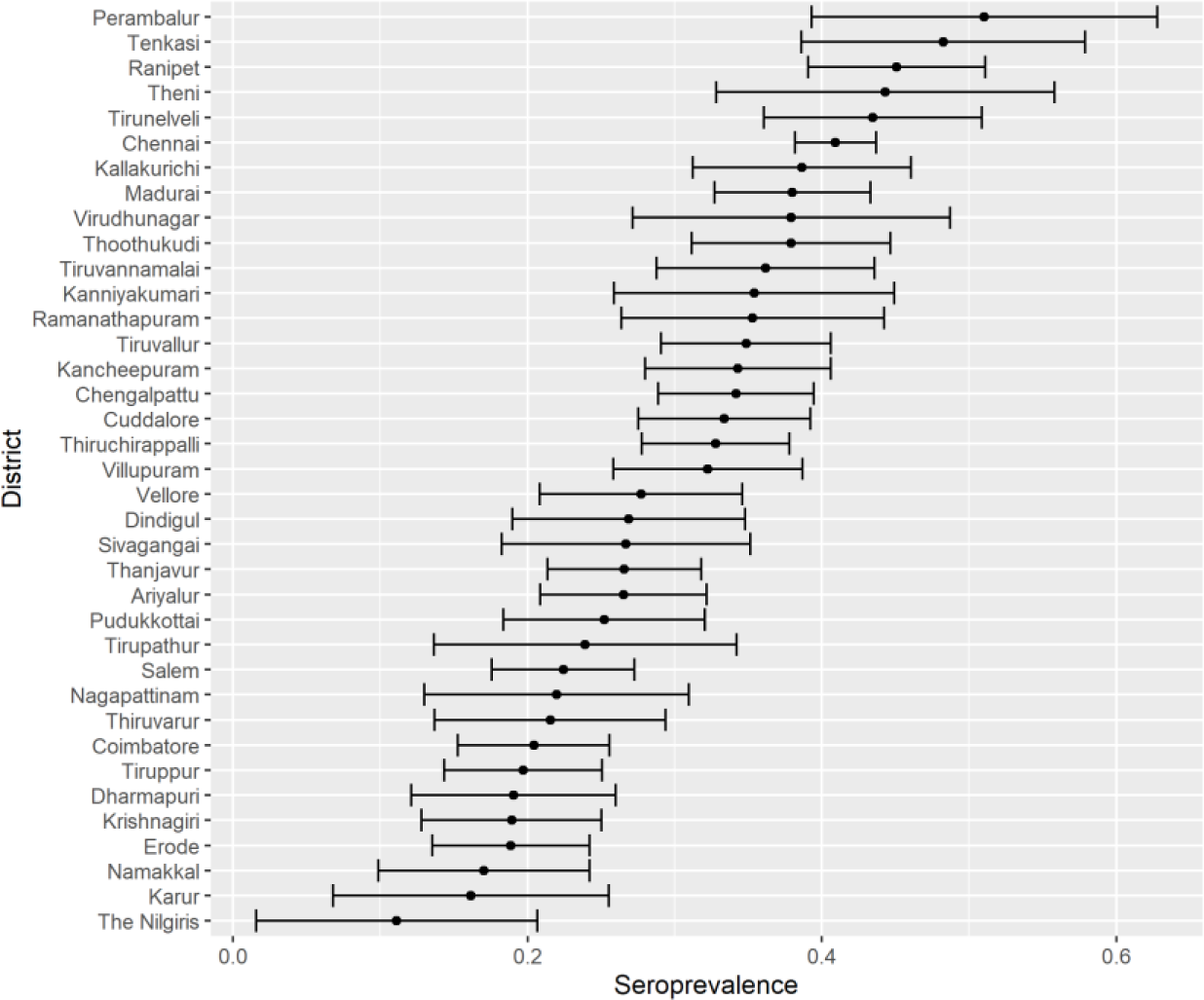
Seroprevalence by district.

**Table 2.** Seroprevalence by type of region, sex, and age.

| **Variable** |  | **Seropre-**<br>**valence** | **CI lower**<br>**bound** | **CI upper**<br>**bound** |
| --- | --- | --- | --- | --- |
| Region | Rural | 0.251 | 0.242 | 0.261 |
|  | Urban | 0.367 | 0.357 | 0.377 |
| Sex | Male | 0.304 | 0.296 | 0.312 |
|  | Female | 0.321 | 0.311 | 0.33 |
| Age | 18-29 | 0.311 | 0.303 | 0.318 |
|  | 30-39 | 0.32 | 0.312 | 0.327 |
|  | 40-49 | 0.333 | 0.325 | 0.34 |
|  | 50-59 | 0.332 | 0.324 | 0.339 |
|  | 60-69 | 0.284 | 0.277 | 0.291 |
|  | 70+ | 0.252 | 0.245 | 0.258 |

Note. Calculated over a sample representative of the entire state.

Seroprevalence is not significantly different across sexes (females, 30.8%; males; 30.3%; p=0.25) (Table 2). Seroprevalence among the elderly (70+: 25.8%) is significantly lower than among the working age populations (age 40-49: 31.6%; p<0.001) or the young (18-29: 30.7%; p<0.001), respectively (Table 2).

The IFR varies substantially across the state, from 0.007% in Perambalur to 0.203% in Chennai (Table 3). The IFR increases with age (ρ=0.8791; p=0.021) and is higher among males (0.11%) than among females (0.04%; p=<0.001) (Table 4).

**Table 3.** Infection fatality rate, undercount of infections, and testing by district.

| District | Deaths | Confirmed cases | Tests conducted | Seroprevalence (%) | Population | IFR (%) | Ratio of actual to confirmed cases |
| --- | --- | --- | --- | --- | --- | --- | --- |
| Ariyalur | 47 | 4191 | 76279 | 26.52% | 754894 | 0.0235% | 48 |
| Chengalpattu | 673 | 40241 | 399263 | 34.19% | 2556244 | 0.0770% | 22 |
| Chennai | 3862 | 207390 | 1350491 | 40.94% | 4646732 | 0.2030% | 9 |
| Coimbatore | 557 | 37932 | 458054 | 20.43% | 3458045 | 0.0789% | 19 |
| Cuddalore | 271 | 22170 | 263947 | 33.37% | 2605914 | 0.0312% | 39 |
| Dharmapuri | 48 | 4954 | 100746 | 19.06% | 1506843 | 0.0167% | 58 |
| Dindigul | 186 | 9465 | 168422 | 26.88% | 2159775 | 0.0320% | 61 |
| Erode | 124 | 8644 | 207777 | 18.88% | 2251744 | 0.0292% | 49 |
| Kallakurichi | 103 | 9802 | 133384 | 38.66% | 1370281 | 0.0194% | 54 |
| Kancheepuram | 385 | 23941 | 328692 | 34.30% | 1166401 | 0.0962% | 17 |
| Kanniyakumari | 246 | 14158 | 226184 | 35.40% | 1870374 | 0.0372% | 47 |
| Karur | 44 | 3664 | 65578 | 16.16% | 1064493 | 0.0256% | 47 |
| Krishnagiri | 106 | 5857 | 68403 | 18.92% | 1883731 | 0.0297% | 61 |
| Madurai | 424 | 18014 | 364893 | 38.00% | 3038252 | 0.0367% | 64 |
| Nagapattinam | 110 | 5959 | 93702 | 21.99% | 1616450 | 0.0309% | 60 |
| Namakkal | 94 | 7845 | 125838 | 17.04% | 1726601 | 0.0320% | 37 |
| Perambalur | 21 | 2010 | 43049 | 51.05% | 565223 | 0.0073% | 144 |
| Pudukkottai | 148 | 10001 | 137332 | 25.21% | 1618345 | 0.0363% | 41 |
| Ramanathapuram | 127 | 5778 | 105173 | 35.30% | 1353445 | 0.0266% | 83 |
| Ranipet | 177 | 14326 | 128022 | 45.09% | 1210277 | 0.0324% | 38 |
| Salem | 425 | 25144 | 377688 | 22.44% | 3482056 | 0.0544% | 31 |
| Sivagangai | 126 | 5580 | 107979 | 26.68% | 1339101 | 0.0353% | 64 |
| Tenkasi | 153 | 7702 | 108215 | 48.24% | 1407627 | 0.0225% | 88 |
| Thanjavur | 221 | 14486 | 257680 | 26.58% | 2405890 | 0.0346% | 44 |
| The Nilgiris | 38 | 5510 | 120113 | 11.12% | 735394 | 0.0465% | 15 |
| Theni | 193 | 15888 | 170530 | 44.33% | 1245899 | 0.0349% | 35 |
| Thiruchirappalli | 168 | 11645 | 196207 | 32.79% | 2722290 | 0.0188% | 77 |
| Thiruvarur | 99 | 8620 | 133411 | 21.56% | 1264277 | 0.0363% | 32 |
| Thoothukudi | 132 | 14391 | 198278 | 37.91% | 1750176 | 0.0199% | 46 |
| Tirunelveli | 208 | 13684 | 198389 | 43.47% | 1665253 | 0.0287% | 53 |
| Tirupathur | 117 | 5843 | 113189 | 23.93% | 1111812 | 0.0440% | 46 |
| Tiruppur | 176 | 10209 | 187803 | 19.71% | 2479052 | 0.0360% | 48 |
| Tiruvallur | 619 | 35320 | 450341 | 34.85% | 3728104 | 0.0476% | 37 |
| Tiruvannamalai | 262 | 16804 | 216551 | 36.18% | 2464875 | 0.0294% | 53 |
| Vellore | 306 | 16854 | 208871 | 27.72% | 1614242 | 0.0684% | 27 |
| Villupuram | 105 | 12712 | 205430 | 32.25% | 2093003 | 0.0156% | 53 |
| Virudhunagar | 221 | 14980 | 266562 | 37.92% | 1942288 | 0.0300% | 49 |

Notes. Death counts are up to 2 days after date of serological sampling. Test (RT-PCR, not serological) and confirmed case counts are up to 7 days before the median date of serological sampling.
Population is from 2011 Census. Ratio of actual cases to confirmed cases uses seroprevalence times the population as the numerator and confirmed cases as the denominator.

**Table 4.** Infection fatality rate by sex and age.

| Age | Sex | Deaths | Seropre-<br>valence | IFR |
| --- | --- | --- | --- | --- |
| 0-17 | Female | 18 |  | N/A |
| 18-29 | Female | 50 | 29.15% | 0.002% |
| 30-39 | Female | 119 | 32.65% | 0.006% |
| 40-49 | Female | 301 | 32.58% | 0.019% |
| 50-59 | Female | 660 | 32.67% | 0.060% |
| 60-69 | Female | 974 | 28.65% | 0.143% |
| 70+ | Female | 1091 | 27.84% | 0.266% |
| 0-17 | Male | 14 |  | N/A |
| 18-29 | Male | 71 | 32.21% | 0.003% |
| 30-39 | Male | 249 | 28.87% | 0.015% |
| 40-49 | Male | 681 | 30.53% | 0.045% |
| 50-59 | Male | 1761 | 31.35% | 0.164% |
| 60-69 | Male | 2491 | 28.81% | 0.380% |
| 70+ | Male | 3235 | 25.28% | 0.923% |

Notes. Death numbers are total up to 2 days after last date of surveillance. IFR for ages <17 is unavailable because this group is excluded from the sero-surveillance sampling.

Ratio of actual cases to confirmed cases ranges from 9 in Chennai to 144 in Perambalur (Table 3). There is a negative and significant (ρ=-0.58; p<0.001) correlation between testing rate and the undercount rate (Figure 3).

**Figure 3.**
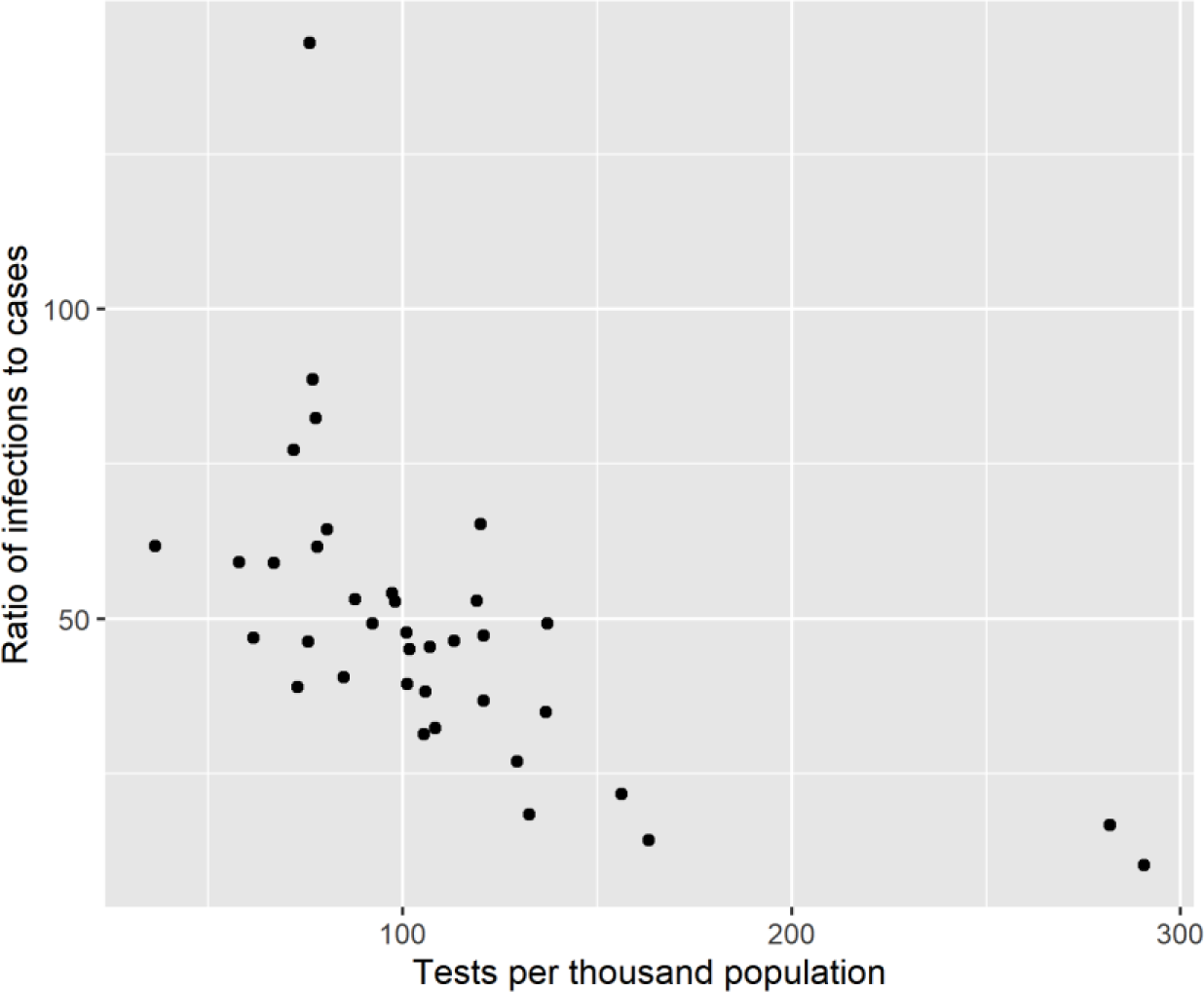
Relationship between rate of undercounting and testing rate. Notes. Each point represents a district. The x-axis presents the number of tests in a district divided by the 2011 Census population in that district. The y-axis presents the ratio of actual cases to confirmed cases. Actual cases are the estimated seroprevalence (%) in the district times its 2011 Census population. The confirmed cases are counts up to 7 days before the median date of serological sampling in the district.

## Discussion

Overall seroprevalence (31.6%) implies that at least 22.7 million persons were infected by November 30, 2020, the last day of serological sampling. Thus, the actual number of infections is roughly 36 times larger than the number of confirmed cases, which totaled 670.392 by 15 October 2020, 7 days before the date the median biosample is collected.

Seroprevalence is highest among working age populations. The lower seroprevalence among the young is not informative about the value of closing schools because the young in our sample were over age 18. Moreover, the difference in seroprevalence among the young and working ages is not significant. The significantly lower seroprevalence among the old is different than what was found in a recent Karnataka seroprevalence study^8^ and suggests that the elderly in Tamil Nadu have been somewhat protected even in multigenerational households.

Higher seroprevalence in urban areas is consistent with higher density in urban districts (Figure 4). It does not seem positively correlated with mobility as measured by average decline in non-residential Google mobility measures.

**Figure 4.**
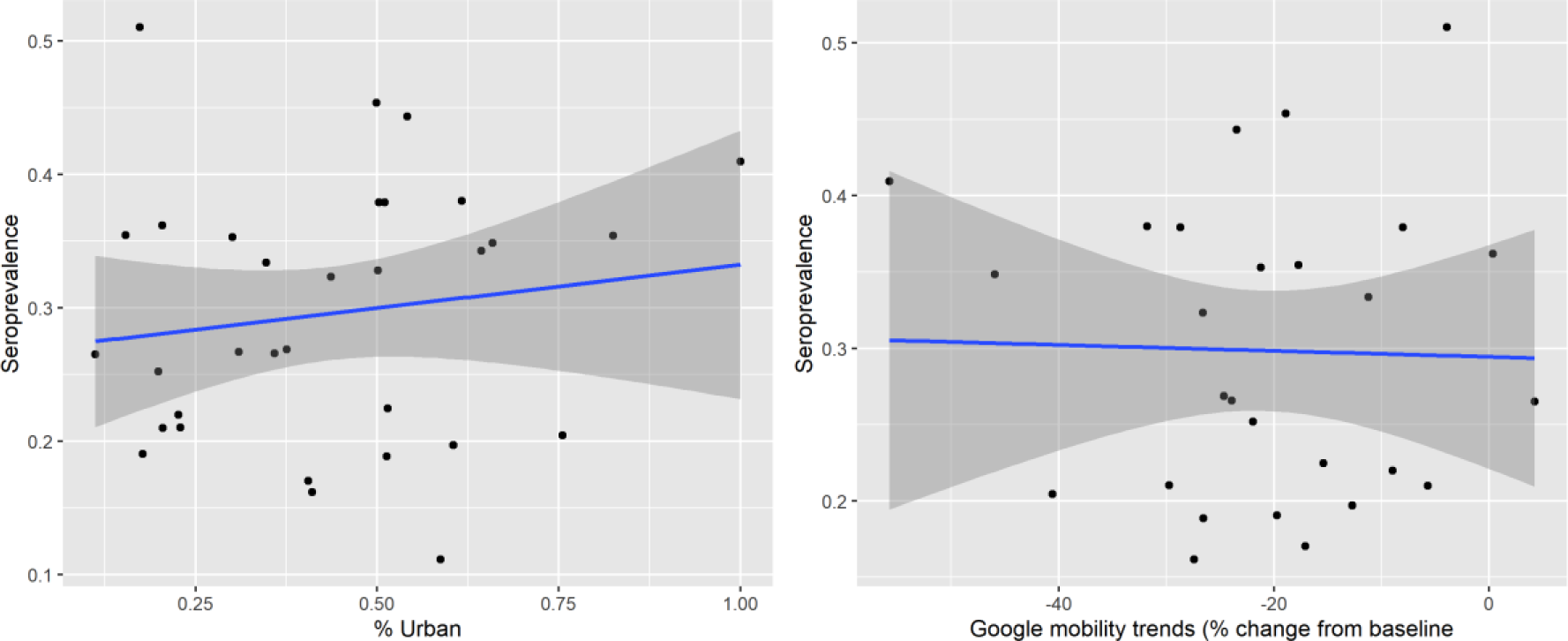
Relationship between seroprevalence, population density, and mobility.

The IFR across the state is 0.052%, nominally lower than that in Karnataka^8^ and Mumbai^9^. The lower IFR is not due to lower infection rate amongst the elderly because IFR is nominally lower even among the elderly. Higher IFR among males is consistent with the literature^10^.

Our study has several limitations. One is that, because antibody concentrations in infected persons decline over time^11^, our estimate of seroprevalence may underestimate the level of prior infection and perhaps natural immunity.

Second, we may underestimate IFR. The number of deaths per million is positively correlated (ρ=0.96; p<0.001) with testing rate per million at the district level (Table 3). Perhaps increasing the testing rate would show greater deaths from SARS-CoV-2.

## SUPPLEMENT

### Methods

#### Sampling

Numerous clusters had less or more than 30 persons sampled. We report those in Table S 1. We retain all samples because it is unclear which samples to drop from clusters with >30 observations.

**Table S1.** Number of samples per cluster.

| Samples<br>per cluster | Number<br>of clusters |
| --- | --- |
| 1 | 4 |
| 4 | 1 |
| 8 | 1 |
| 10 | 1 |
| 11 | 1 |
| 20 | 1 |
| 24 | 1 |
| 26 | 1 |
| 28 | 10 |
| 29 | 31 |
| 30 | 818 |
| 31 | 16 |
| 32 | 2 |
| 36 | 1 |

#### Sample size

Sampling 300 per million would lead to different sample sizes per district. Because some clusters yielded less than 30 persons per cluster, the study produced just 292 samples per 1 million population. The following table presents the sample size obtained and the resulting minimum detectable effect in each district. These are reported in Table S 2.

**Table S2.** Sample size obtained per district and implied minimum detectable effect.

| District | Sample size | MDE | District | Sample size | MDE |
| --- | --- | --- | --- | --- | --- |
| Ariyalur | 270 | 0.119 | Ramanathapuram | 480 | 0.089 |
| Chengalpattu | 720 | 0.073 | Ranipet | 420 | 0.096 |
| Chennai | 3613 | 0.033 | Salem | 1260 | 0.055 |
| Coimbatore | 1182 | 0.057 | Sivagangai | 450 | 0.092 |
| Cuddalore | 870 | 0.066 | Tenkasi | 420 | 0.096 |
| Dharmapuri | 567 | 0.082 | Thanjavur | 834 | 0.068 |
| Dindigul | 717 | 0.073 | The Nilgiris | 223 | 0.131 |
| Erode | 745 | 0.072 | Theni | 420 | 0.096 |
| Kallakurichi | 596 | 0.080 | Thiruchirappalli | 957 | 0.063 |
| Kancheepuram | 480 | 0.089 | Thiruvarur | 420 | 0.096 |
| Kanniyakumari | 659 | 0.076 | Thoothukudi | 547 | 0.084 |
| Karur | 390 | 0.099 | Tirunelveli | 630 | 0.078 |
| Krishnagiri | 690 | 0.075 | Tirupathur | 329 | 0.108 |
| Madurai | 1140 | 0.058 | Tiruppur | 740 | 0.072 |
| Mayiladuthurai | 320 | 0.110 | Tiruvallur | 745 | 0.072 |
| Nagapattinam | 209 | 0.136 | Tiruvannamalai | 810 | 0.069 |
| Namakkal | 600 | 0.080 | Vellore | 592 | 0.081 |
| Perambalur | 209 | 0.136 | Villupuram | 594 | 0.080 |
| Pudukkottai | 599 | 0.080 | Virudhunagar | 688 | 0.075 |

Note. MDE is calculated assuming a prevalence of 0.5, confidence level of 95%, and a design effect of 2.

Assuming a design effect of 2, the implied minimum detectable effect (MDE) per district varies from 3.3 (Chennai) to 13.6 (Nagapattinam) percentage points.

#### Sample

Suspected or confirmed current or prior COVID-19 infection was not an exclusion criterion. If a participant was currently receiving medical care for COVID-19, a family member or proxy was used to complete the questionnaire on the participant’s behalf; however, the blood sample was taken from the participant.

#### Data collection

Blood was collected in EDTA vacutainers. Serum was isolated and stored in Eppendorf tubes. Serum was analyzed using either of two chemiluminescent immunoassay (CLIA) kits.

The first kit was the iFlash-SARS-CoV-2 IgG kit from Shenzhen YHLO Biotech. Per the manufacturer, it has a sensitivity of 95.9% (95% CI: 93.3-97.5%) and specificity of 95.7% (95% CI: 92.5-97.6%)^4^. Independent analysis estimated a sensitivity of 93% (95% CI: 84.3–97.7%) and specificity of 92.9% (95% CI: 85.3–97.4%)^12^.

The second kit was the Vitros anti-SARS-CoV-2 IgG CLIA from Ortho-Clinical Diagnostics. Per the manufacturer it has 90% sensitivity (95% CI: 76.3-97.2%) and 100% specificity (95% CI: 99.1–100.0%)^5^. FDA evaluation suggests it has 100% sensitivity (95% CI: 88.7-100%) and 100% specificity (95% CI: 95.4-100%)^13^. Independent analysis estimated that it has a sensitivity of 98.8% (95% CI: 92.9-100%) and specificity of 97.3% (95% CI: 85-100%)^14^.

All the samples in a district are analyzed using the same kit, with the exception of the Chennai. In Chennai 2 HUDs used 1 kit, one used the other. Table S 3 reports the test kit used in each district.

**Table S3.** Test kit used in each district.

| **District** | **Type of Kit** | **District** | **Type of Kit** |
| --- | --- | --- | --- |
| Ariyalur | CPC Kit | Ramanathapuram | CPC Kit |
| Chengalpattu | Ortho Kit | Ranipet | Ortho Kit |
| Chennai | CPC & Ortho kits* | Salem | Ortho Kit |
| Coimbatore | Ortho Kit | Sivagangai | CPC Kit |
| Cuddalore | CPC Kit | Tenkasi | CPC Kit |
| Dharmapuri | Ortho Kit | Thanjavur | CPC Kit |
| Dindigul | CPC Kit | The Nilgiris | Ortho Kit |
| Erode | Ortho Kit | Theni | CPC Kit |
| Kallakurichi | CPC Kit | Thiruchirappalli | CPC Kit |
| Kancheepuram | Ortho Kit | Thiruvarur | CPC Kit |
| Kanniyakumari | CPC Kit | Thoothukudi | CPC Kit |
| Karur | CPC Kit | Tirunelveli | CPC Kit |
| Krishnagiri | Ortho Kit | Tirupathur | Ortho Kit |
| Madurai | CPC Kit | Tiruppur | Ortho Kit |
| Mayiladuthurai | CPC Kit | Tiruvallur | Ortho Kit |
| Nagapattinam | CPC Kit | Tiruvannamalai | Ortho Kit |
| Namakkal | Ortho Kit | Vellore | Ortho Kit |
| Perambalur | CPC Kit | Villupuram | CPC Kit |
| Pudukkottai | CPC Kit | Virudhunagar | CPC Kit |

Notes. 33 out of 122 clusters in Chennai used the CPC test kit.

#### Statistical analysis

Nagapattinam district was split into Nagapattinam and Mayiladuthurai districts in March 2020, after the state started reported data on confirmed cases but before we conducted our serological survey. We aggregate these two districts together in our estimates of seropositivity and seroprevalence.

In Chennai, we do not have the population by HUDs. Since the samples were drawn proportional to population, we divide the district population across the HUDs in proportion to the sample size.

When estimating our district-level seroprevalence, the weights for our regression analysis employ data from the 2011 Census for the population in each age x gender category in each district. We estimate the sampling probability for demographic group (age category x sex) as the number of observations in that group in the sample in a district divided by the census population in that group in a district.

When estimating our urban- and rural-level seroprevalence, the weights for our regression analysis employ data from the 2011 Census for the population in each urban/rural category in each district. We estimate the sampling probability for urban/rural group as the number of observations in that group in the sample in a district divided by the census population in that group in a district.

We calculate the sampling probabilities for each regression observation at the level of 2011-defined districts (of which there are 32) rather than the 2020-defined districts (of which there are 38), HUDs or clusters because the population is available only at the level of the old 32 districts. Likewise, we calculate district weights when we aggregate estimates across districts using the 32, 2001 districts. The 38, 2020 districts are all the same or bifurcations of the 32, 2011 districts. Fortunately, in all bifurcated districts, the same kit was used. Therefore, we can combine all bifurcated districts into older 2011 districts for purposes of calculating sampling probabilities in regression analyses or weights when aggregating estimates.

### Results

Table S 4 provides the data behind Figure 2, which reports seroprevalence by district. Table S 5 reports seroprevalence by district and demographics.

**Table S4.**
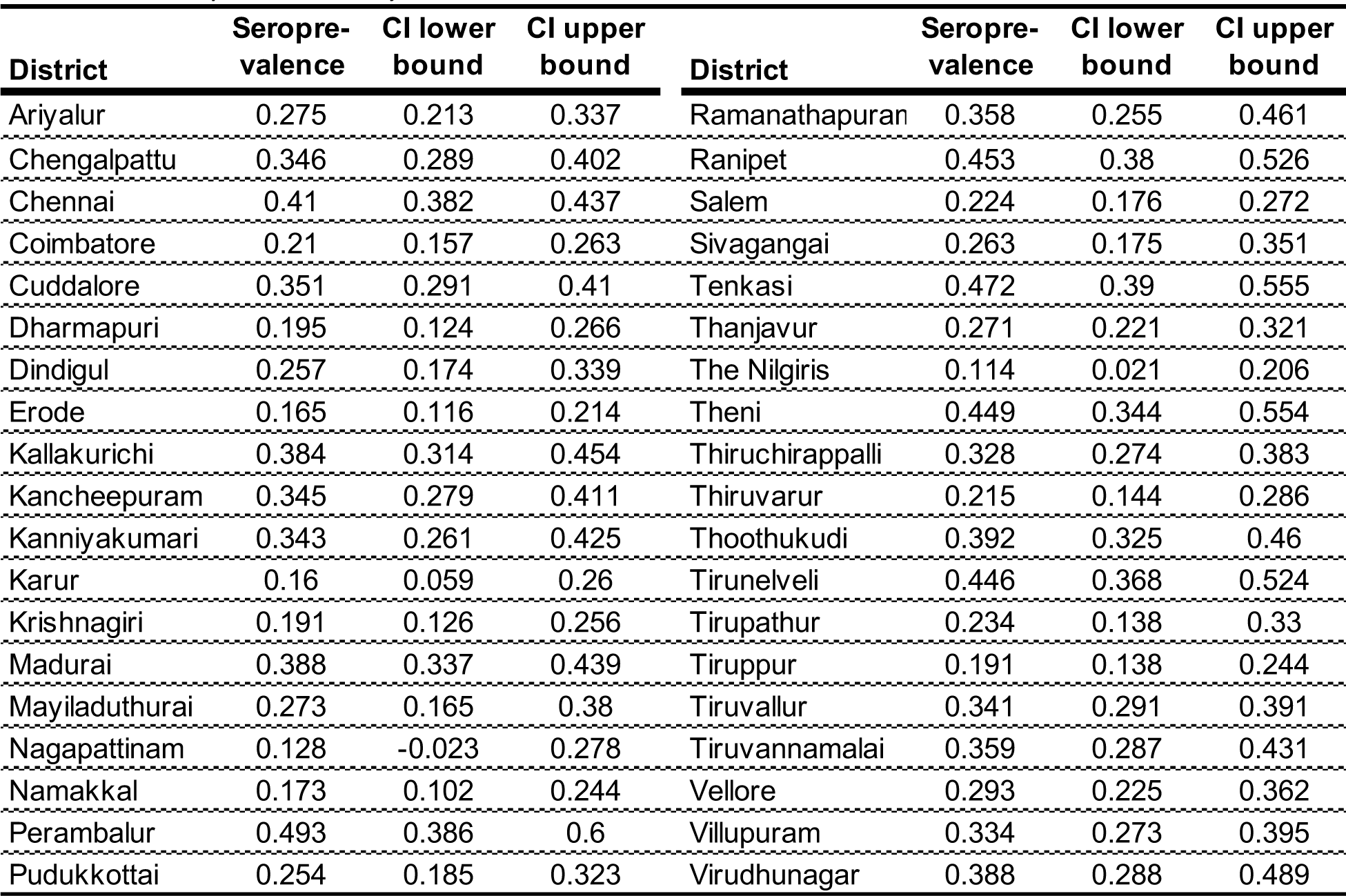
Seroprevalence by district.

**Table S5.**
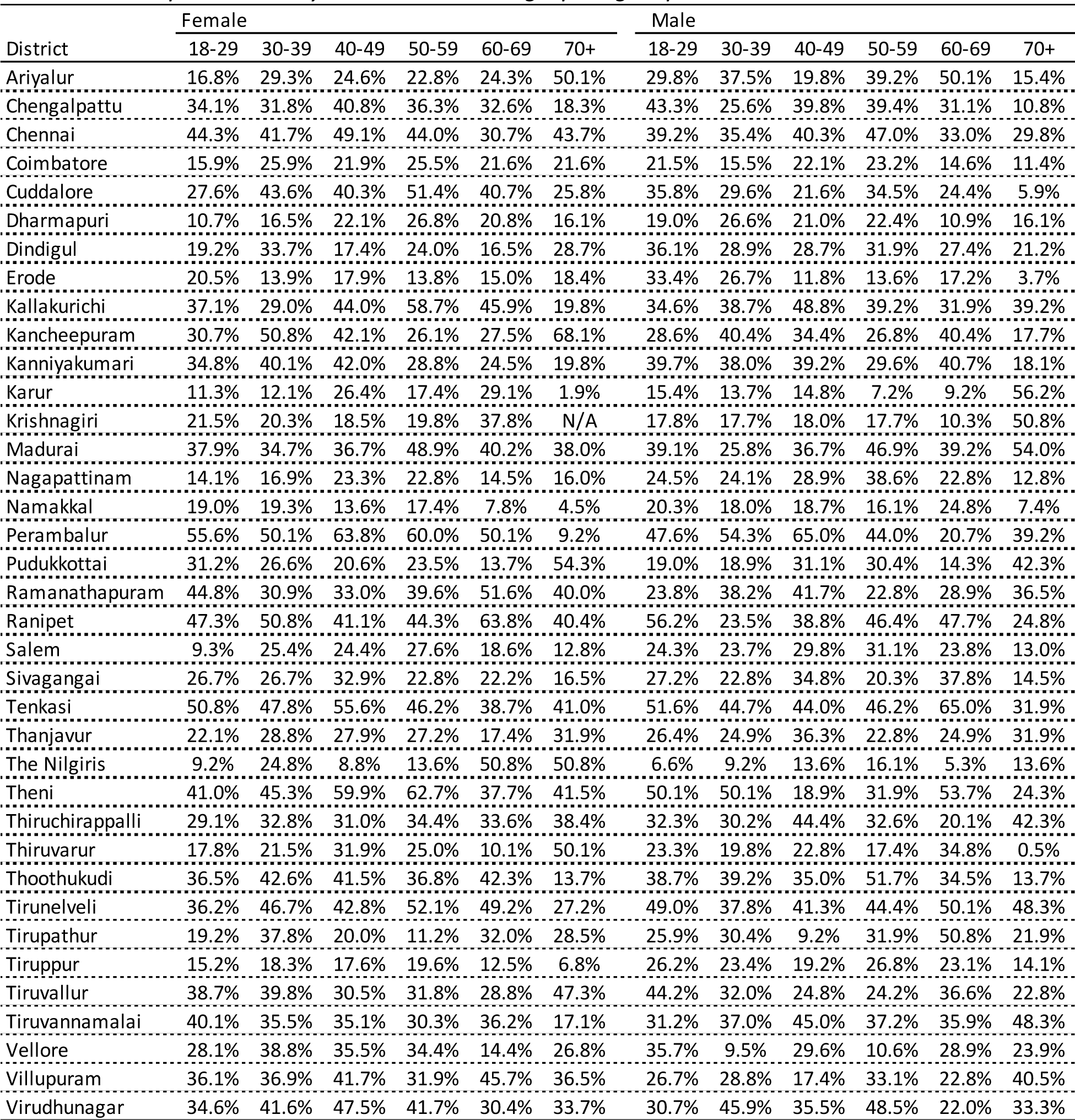
Seroprevalence by district and demographic group.

**Figure S1.**
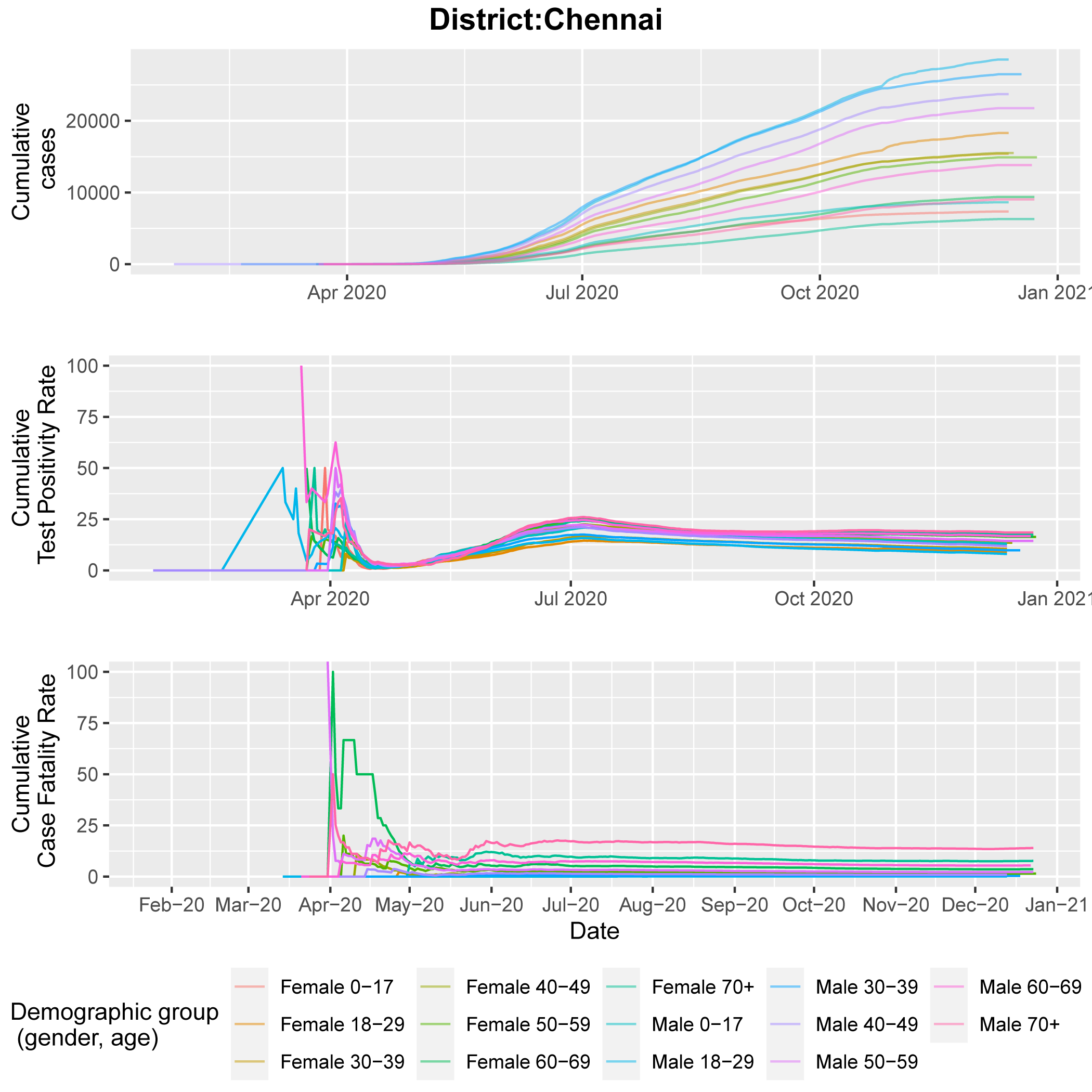
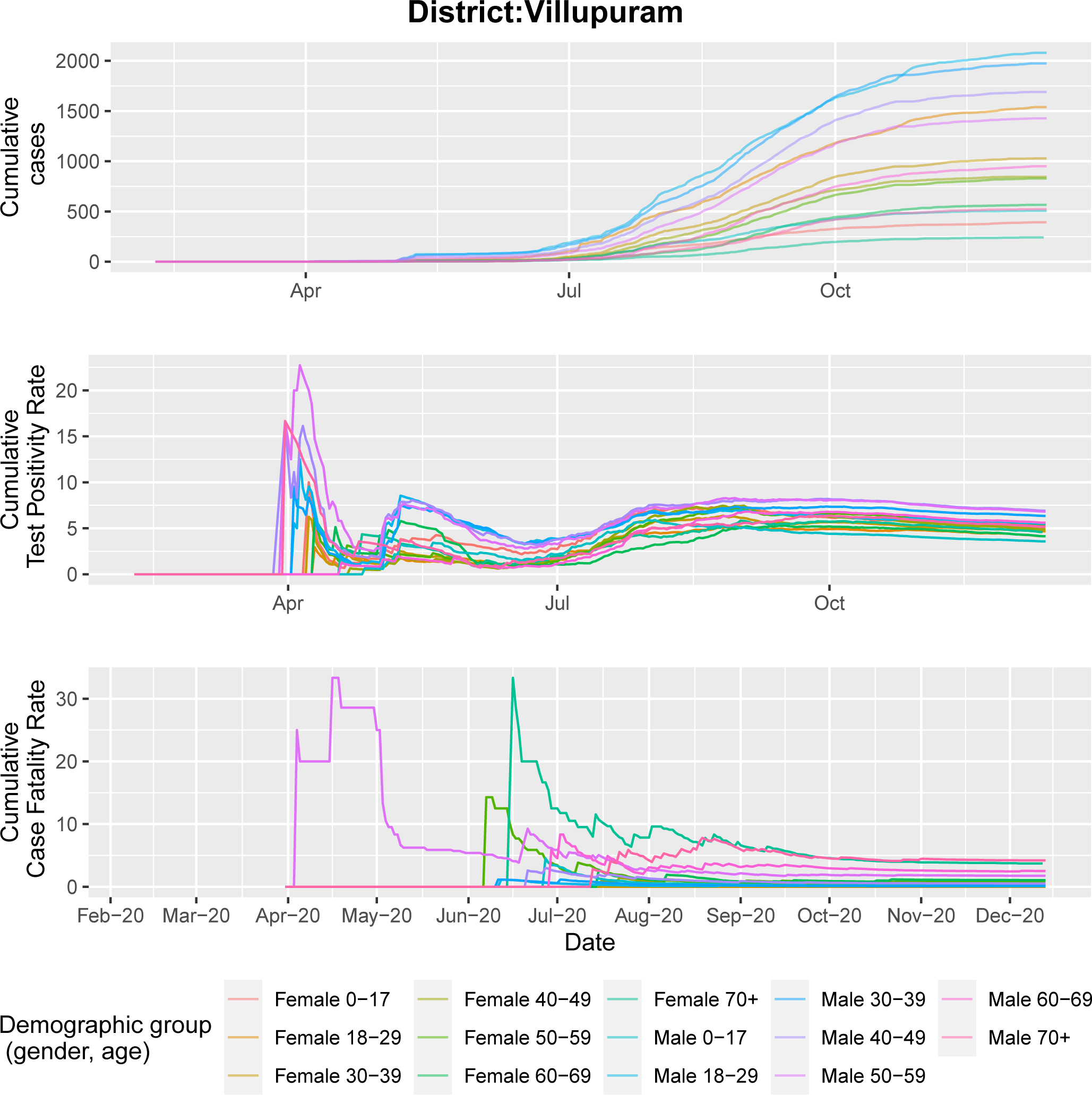
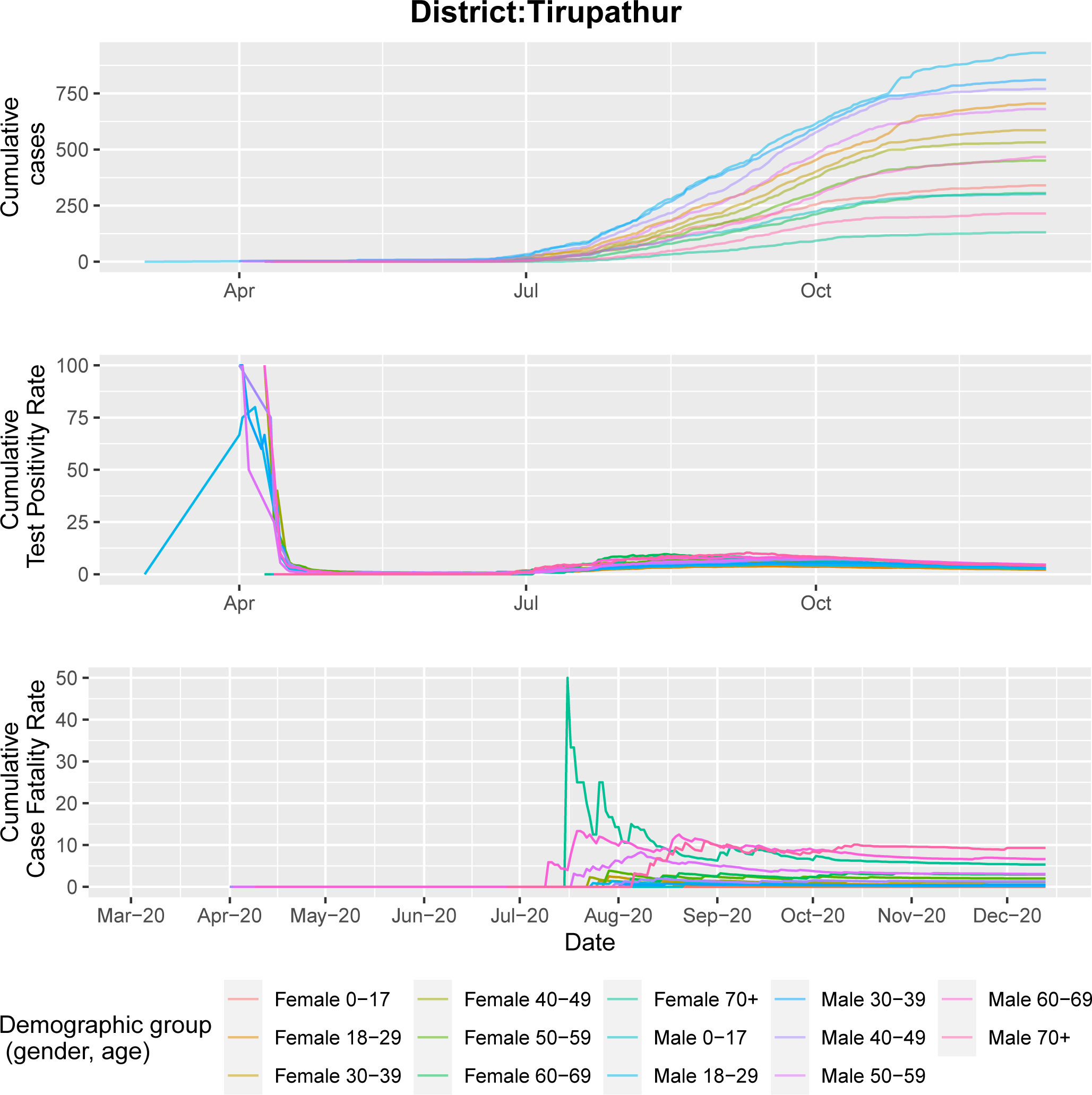
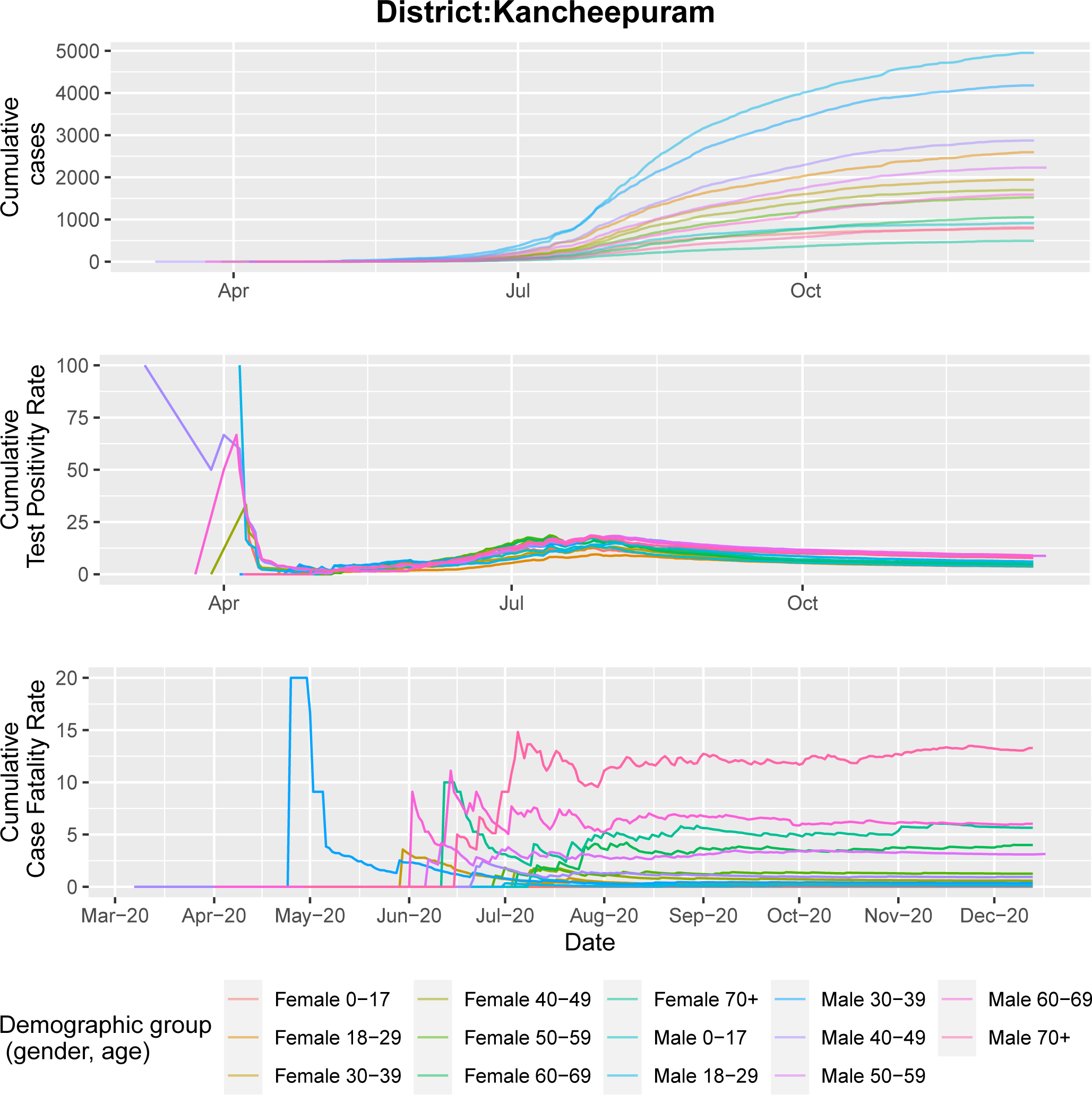
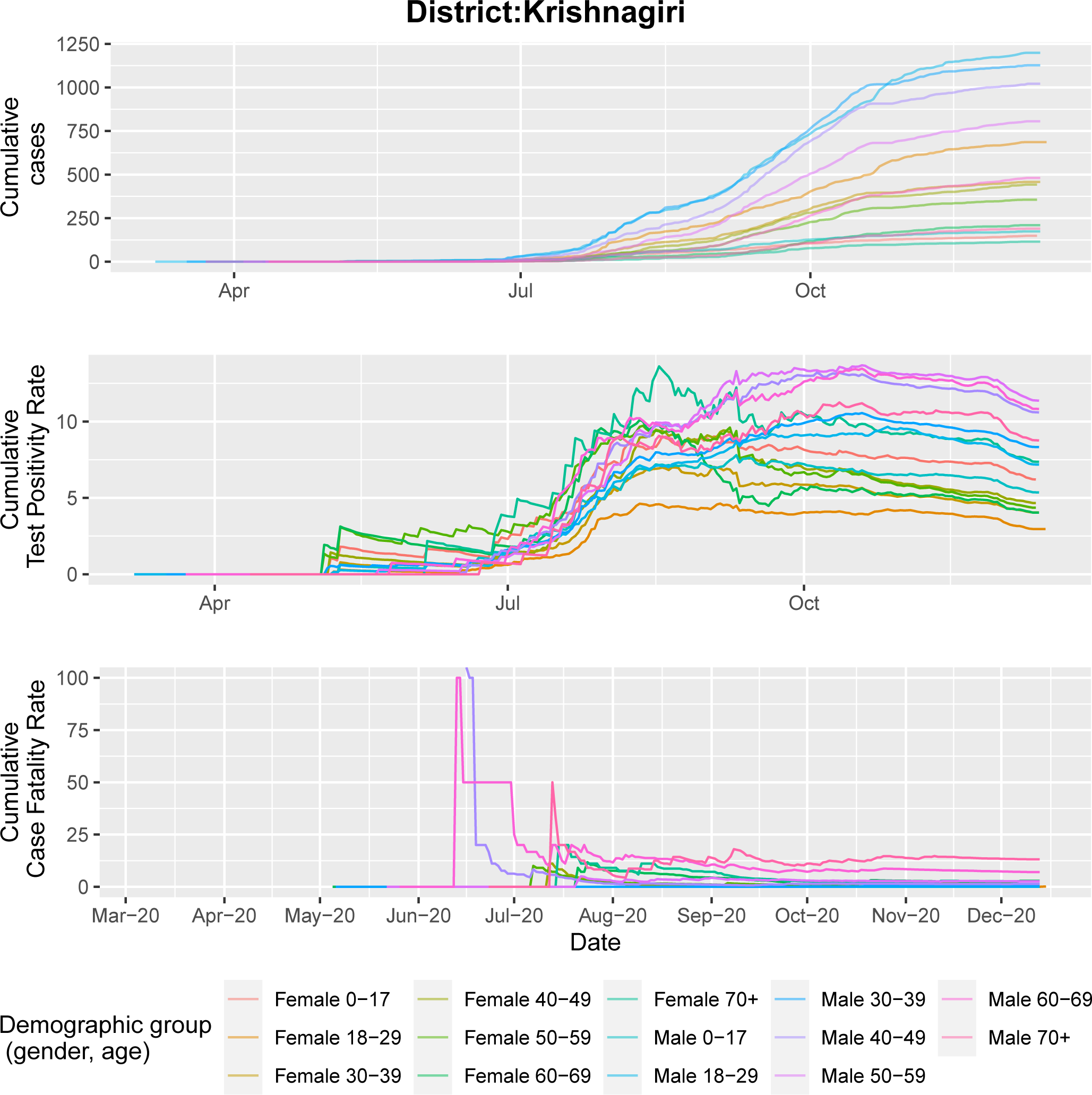
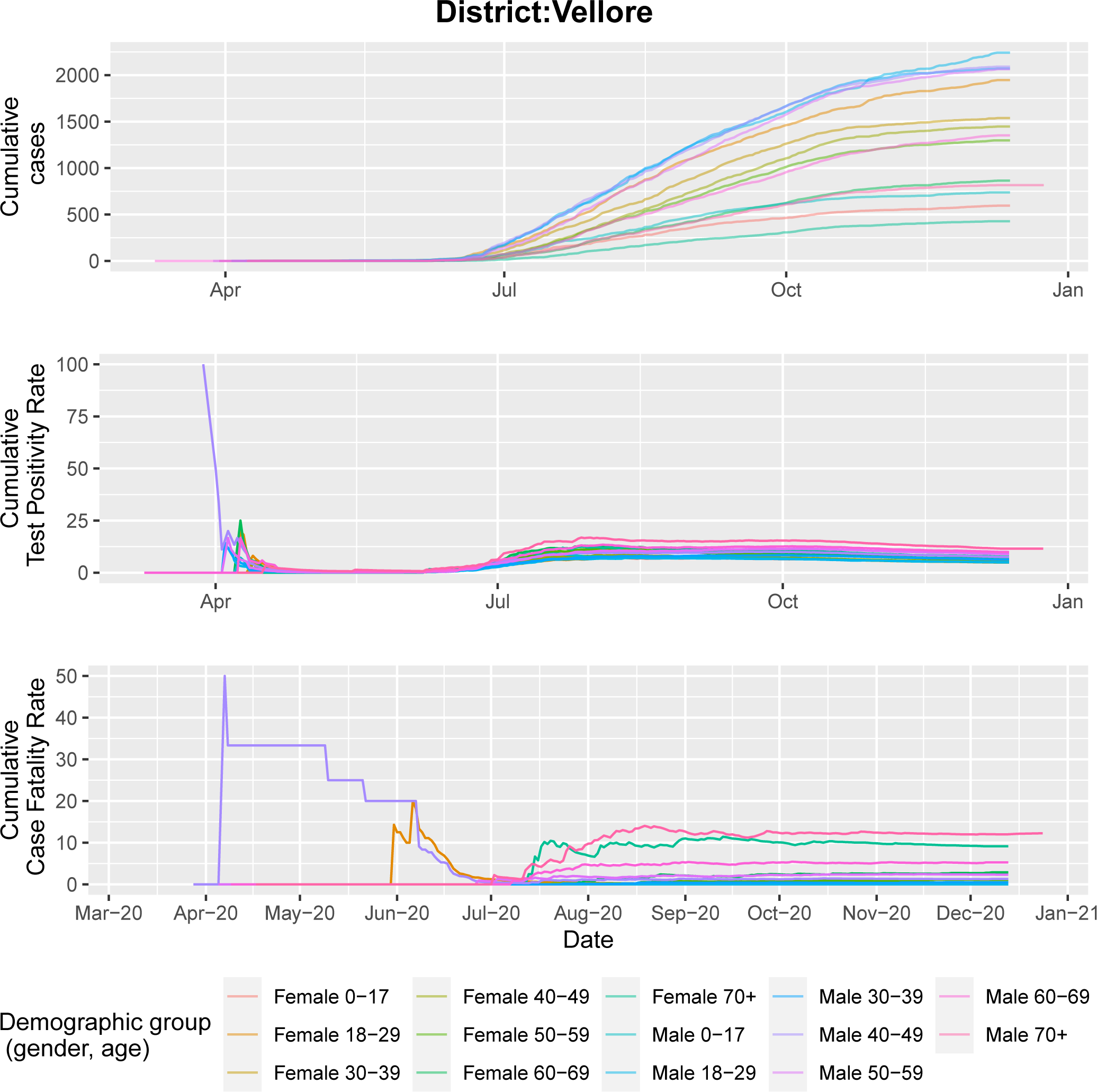
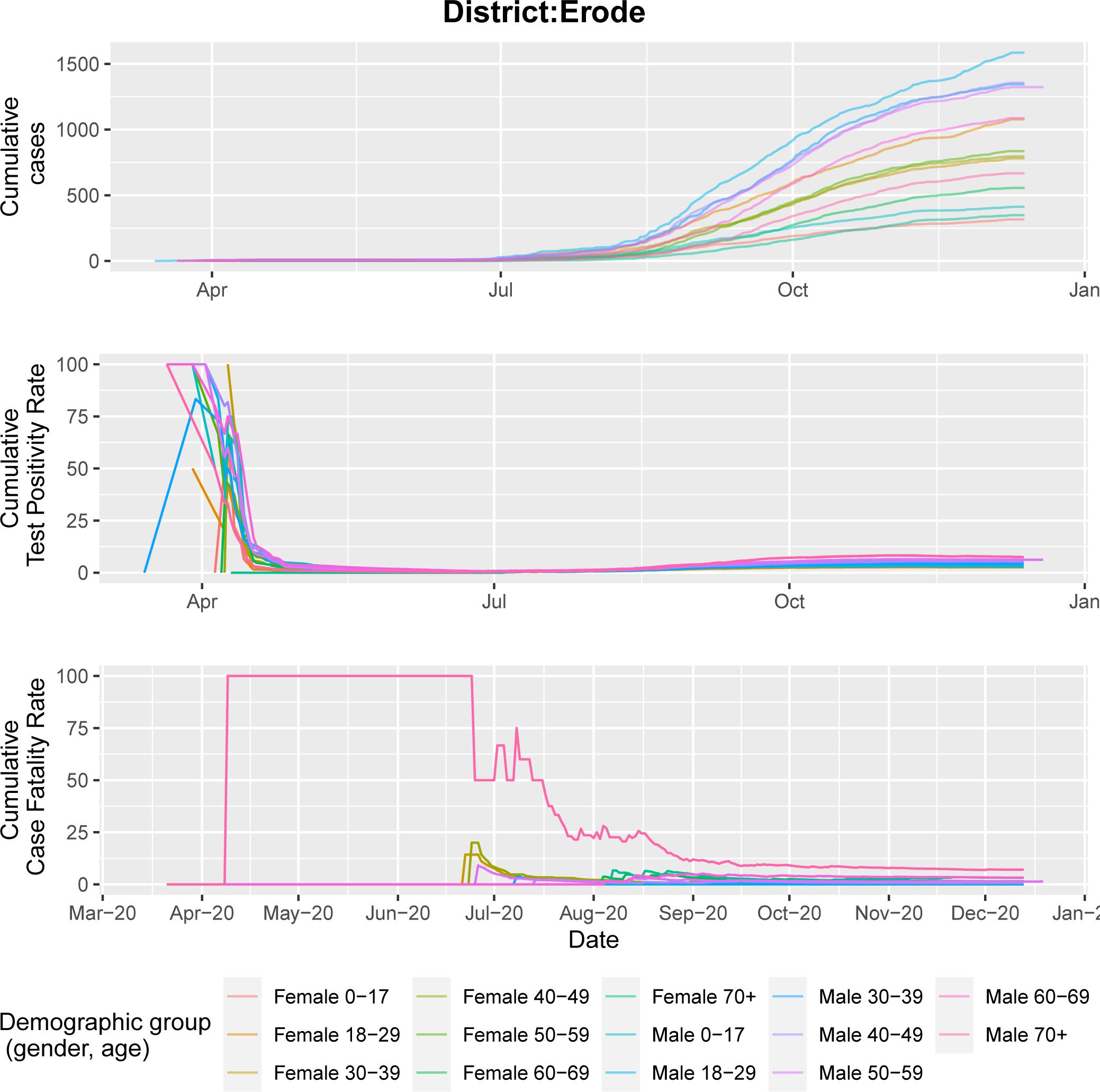
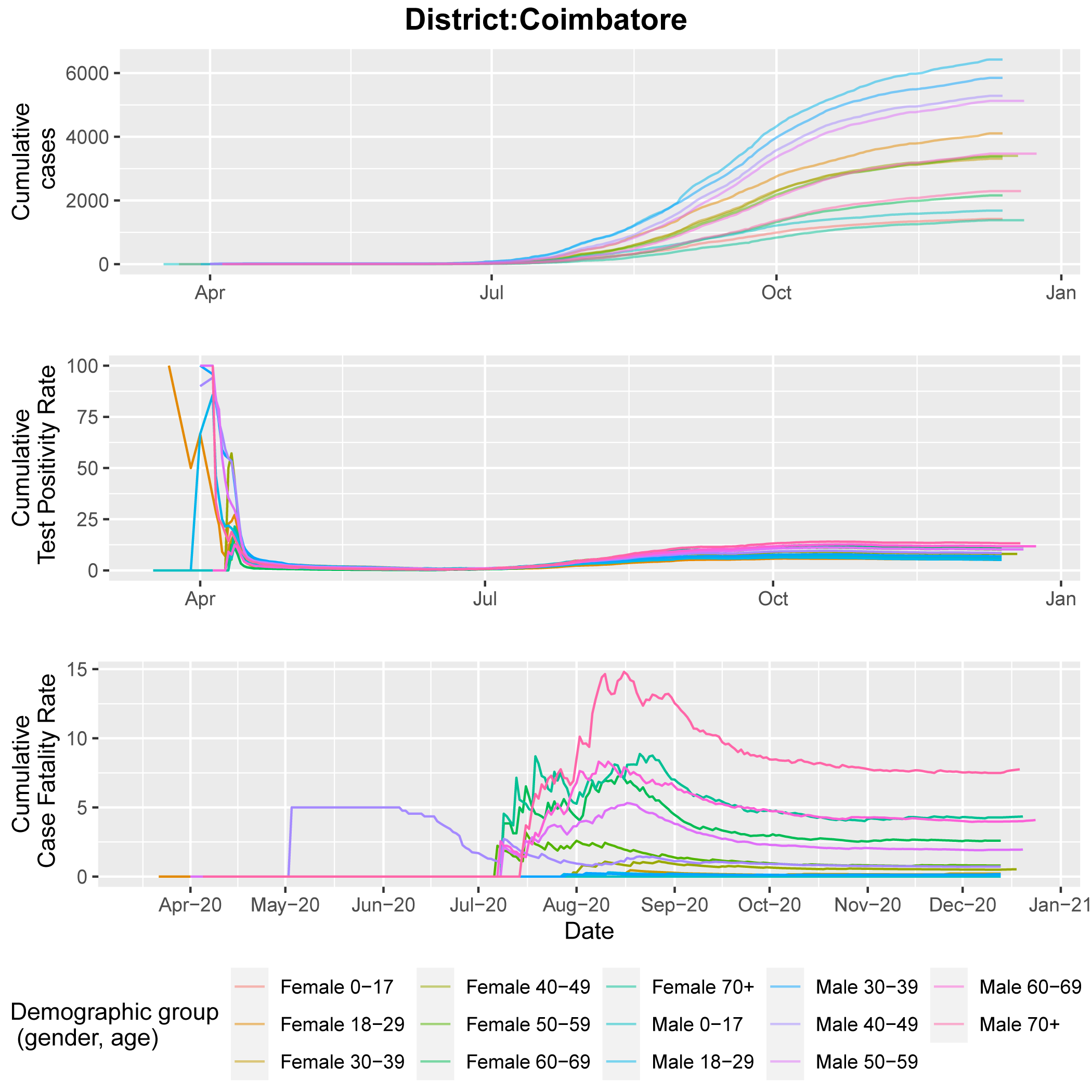
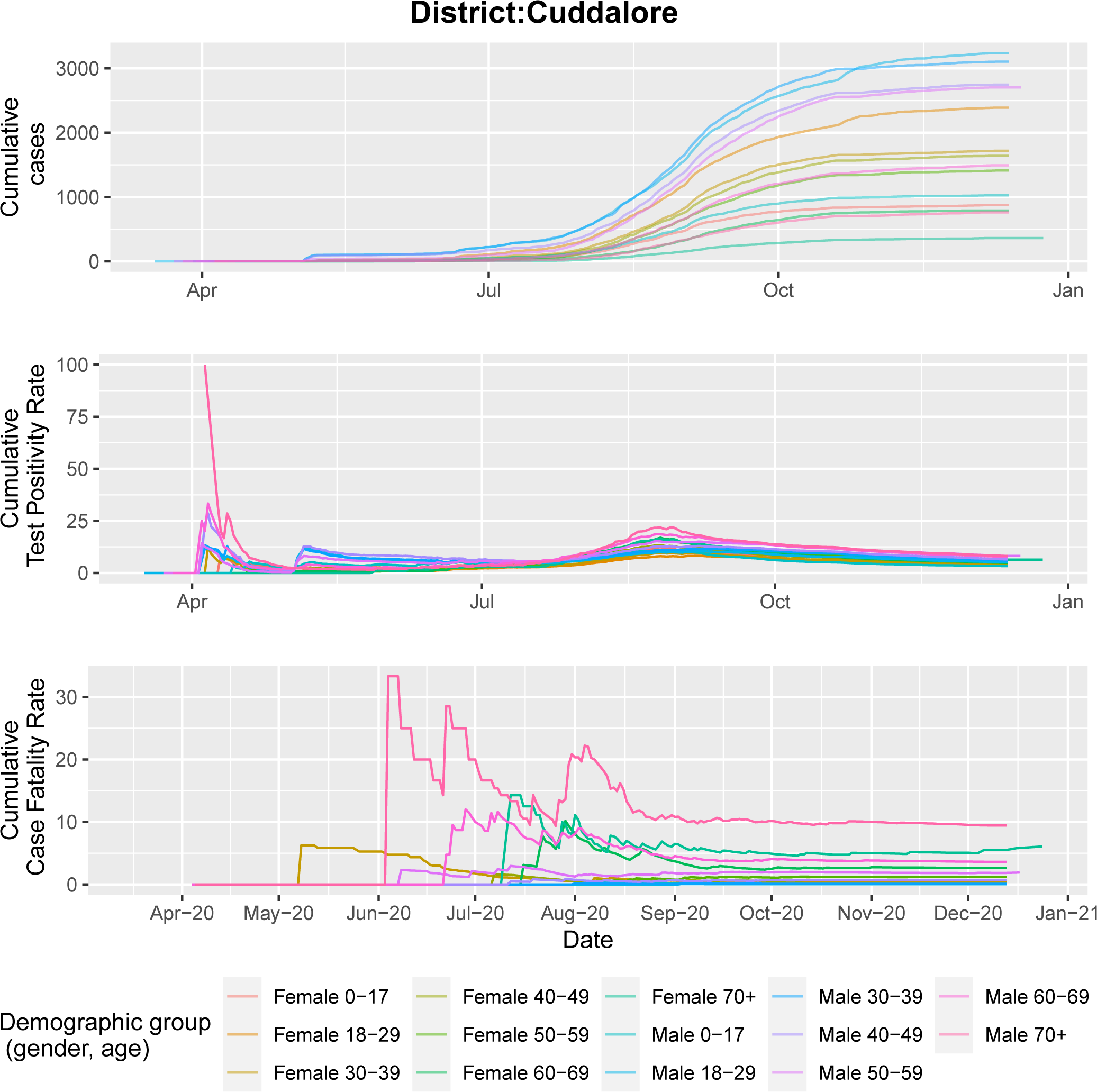
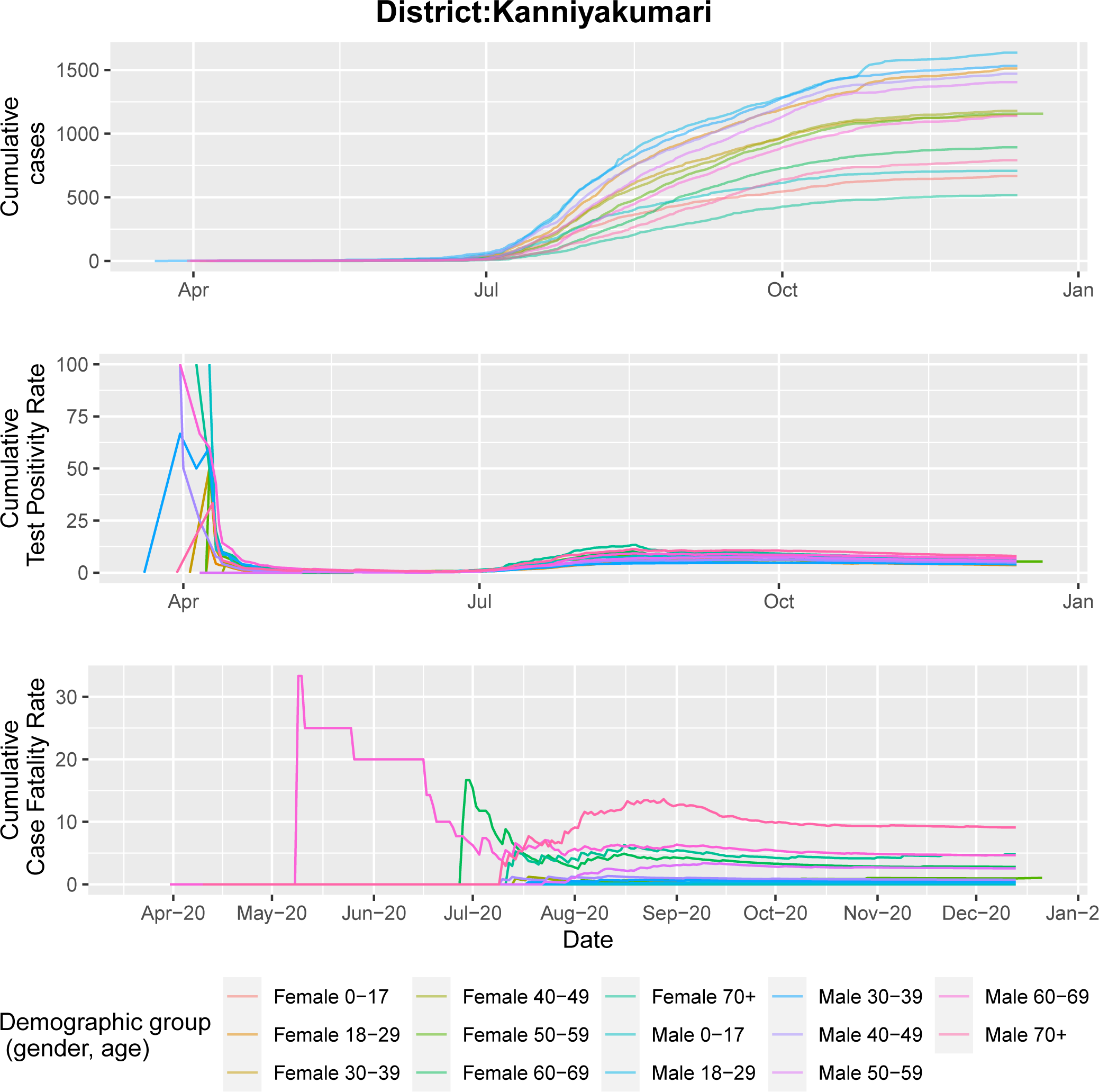
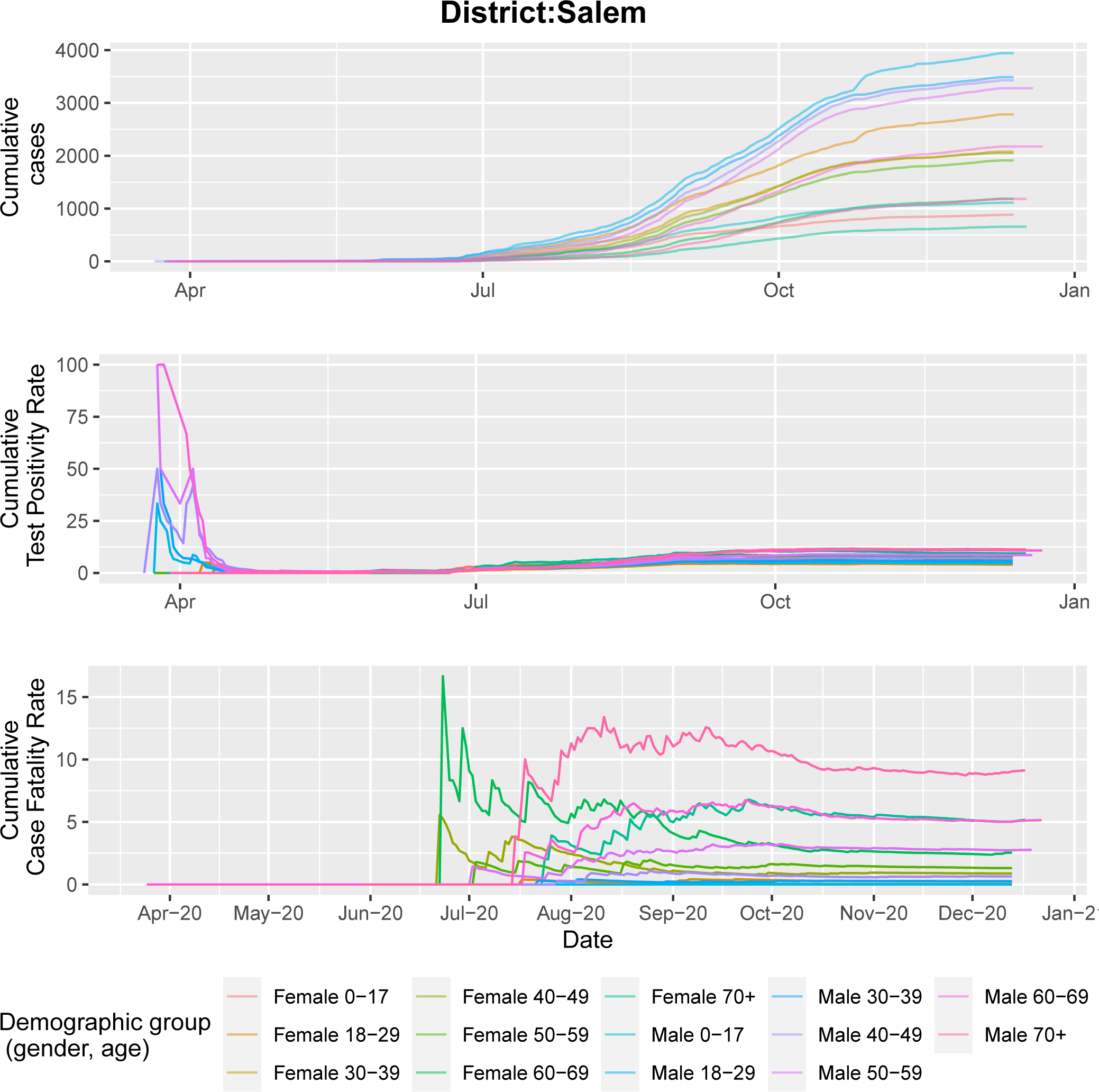
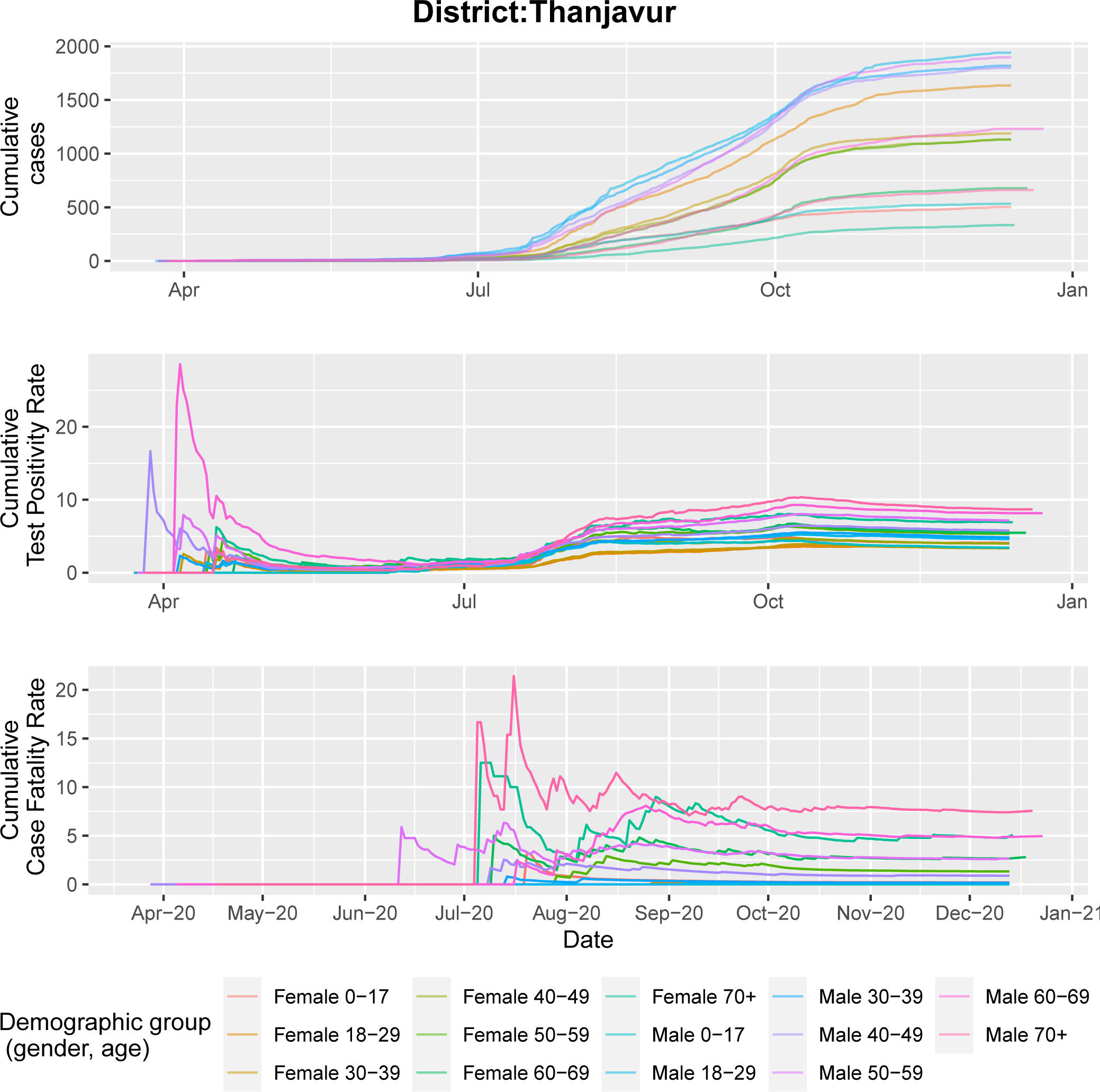
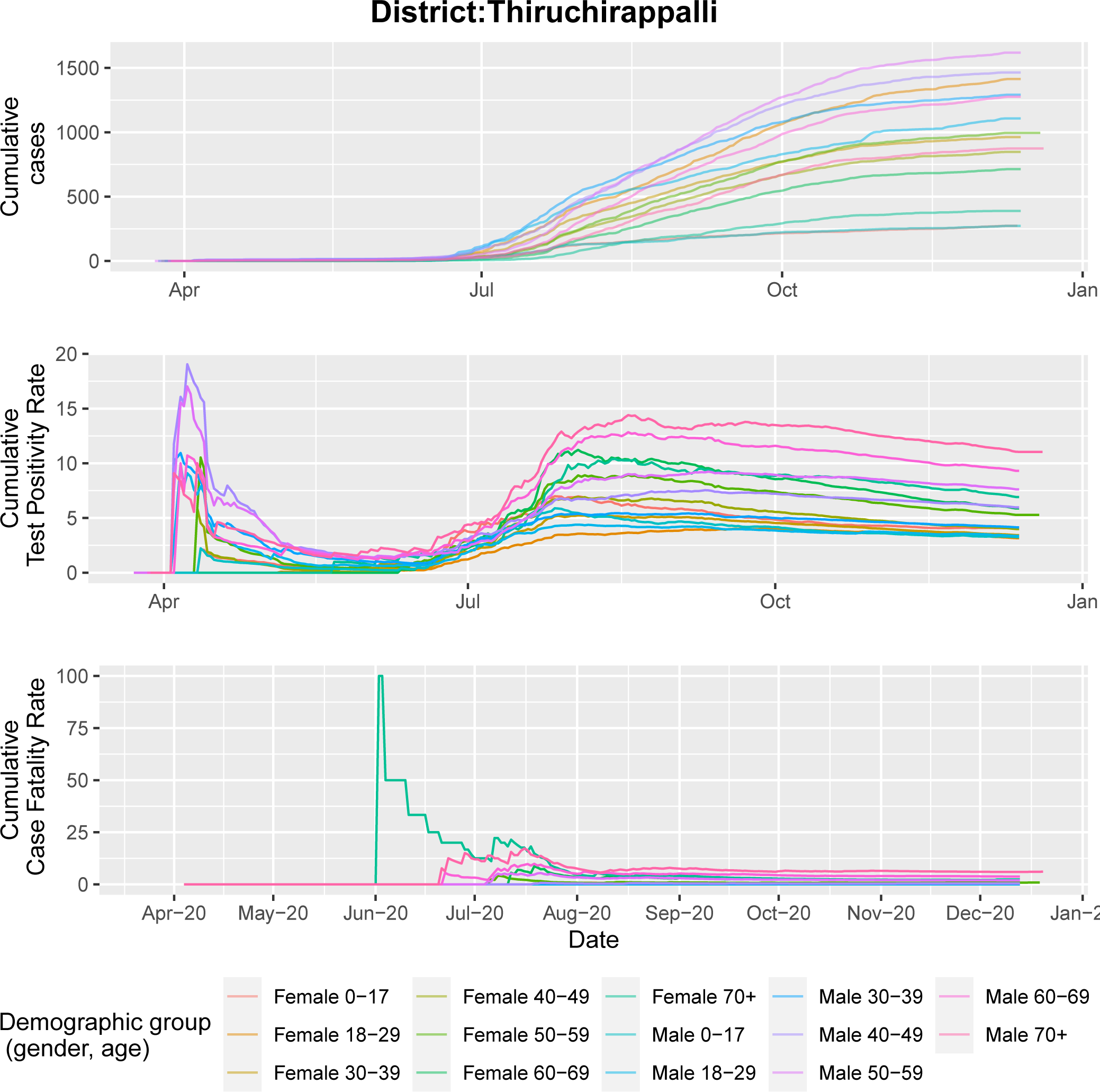
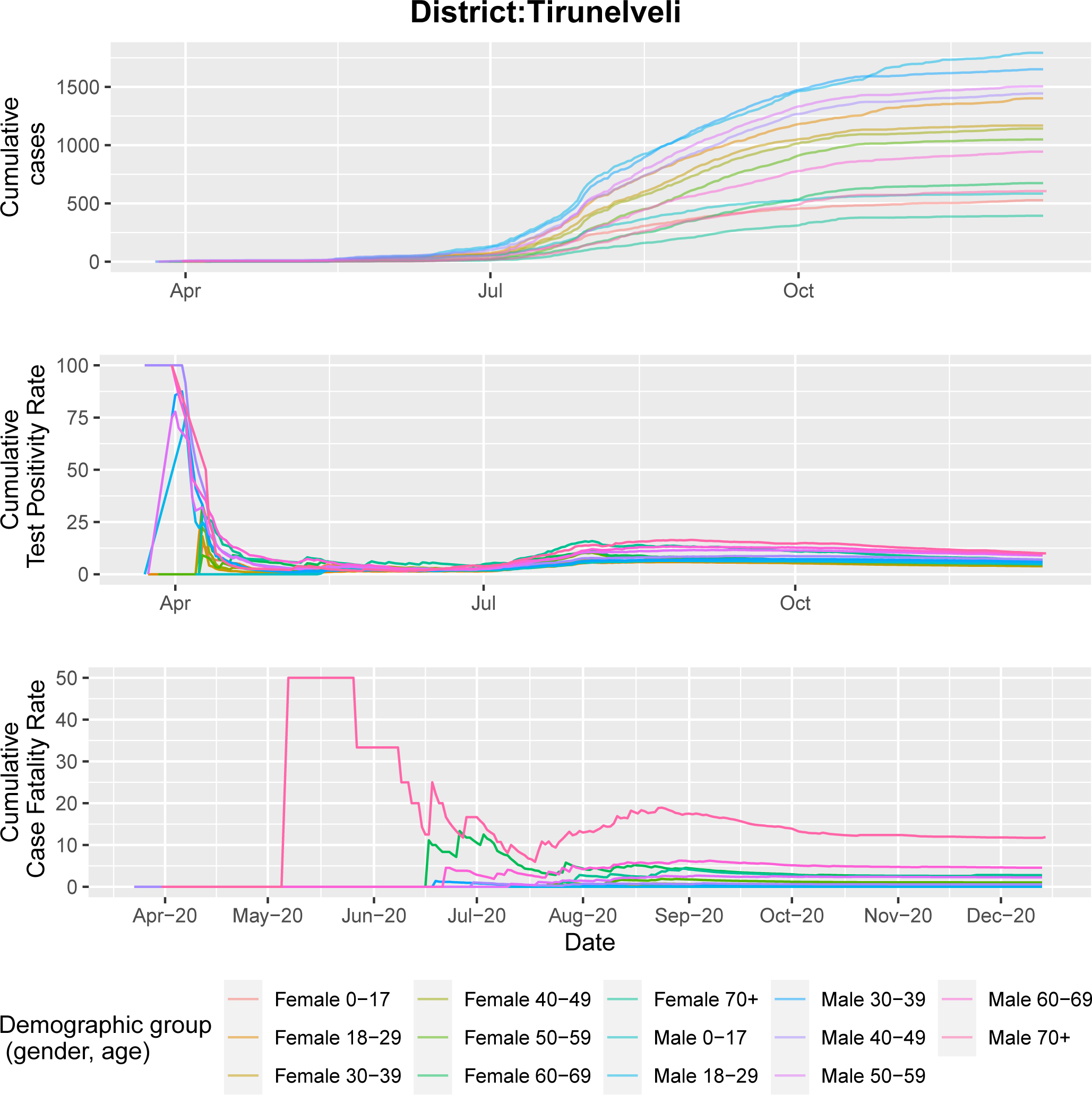
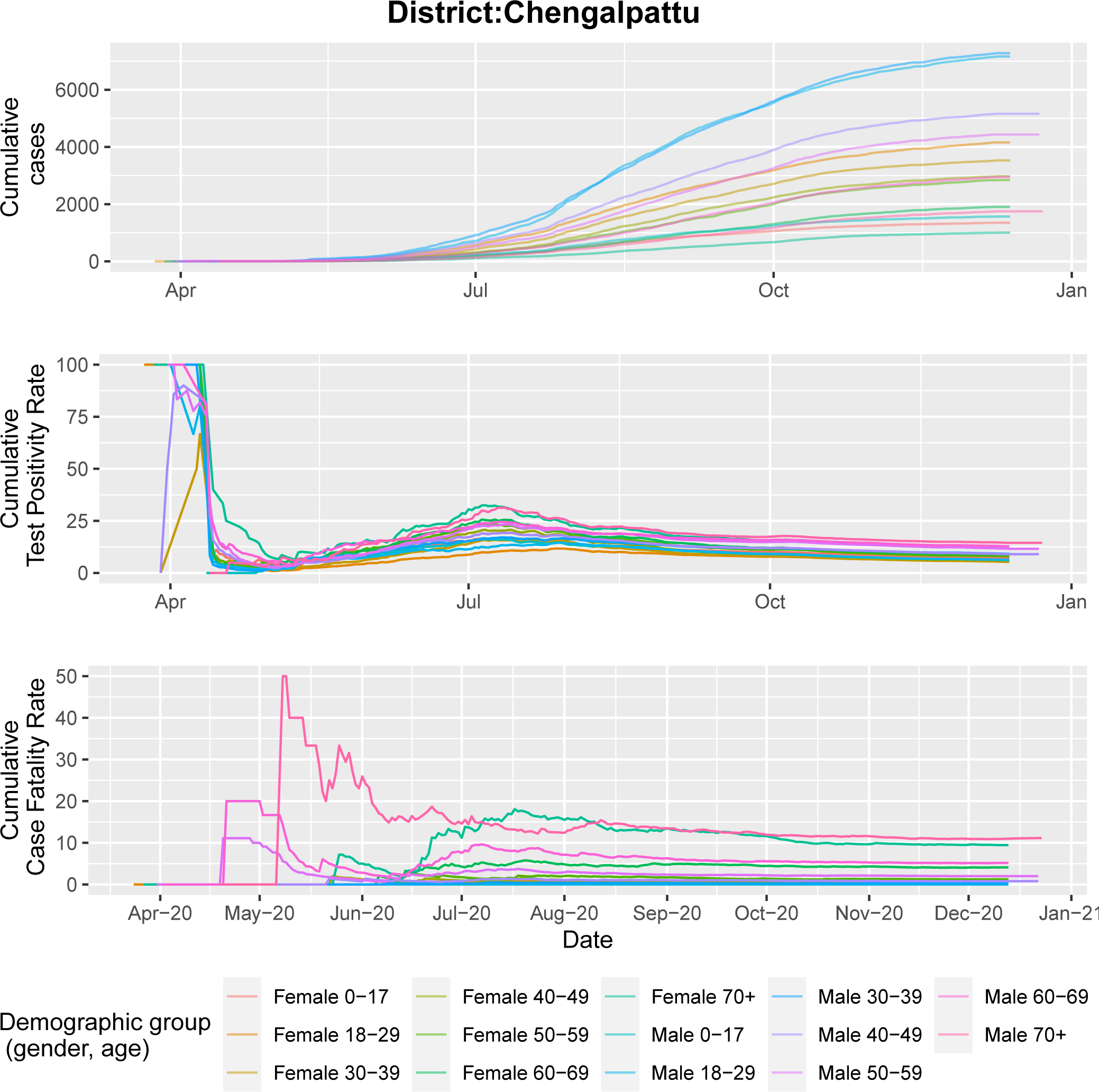
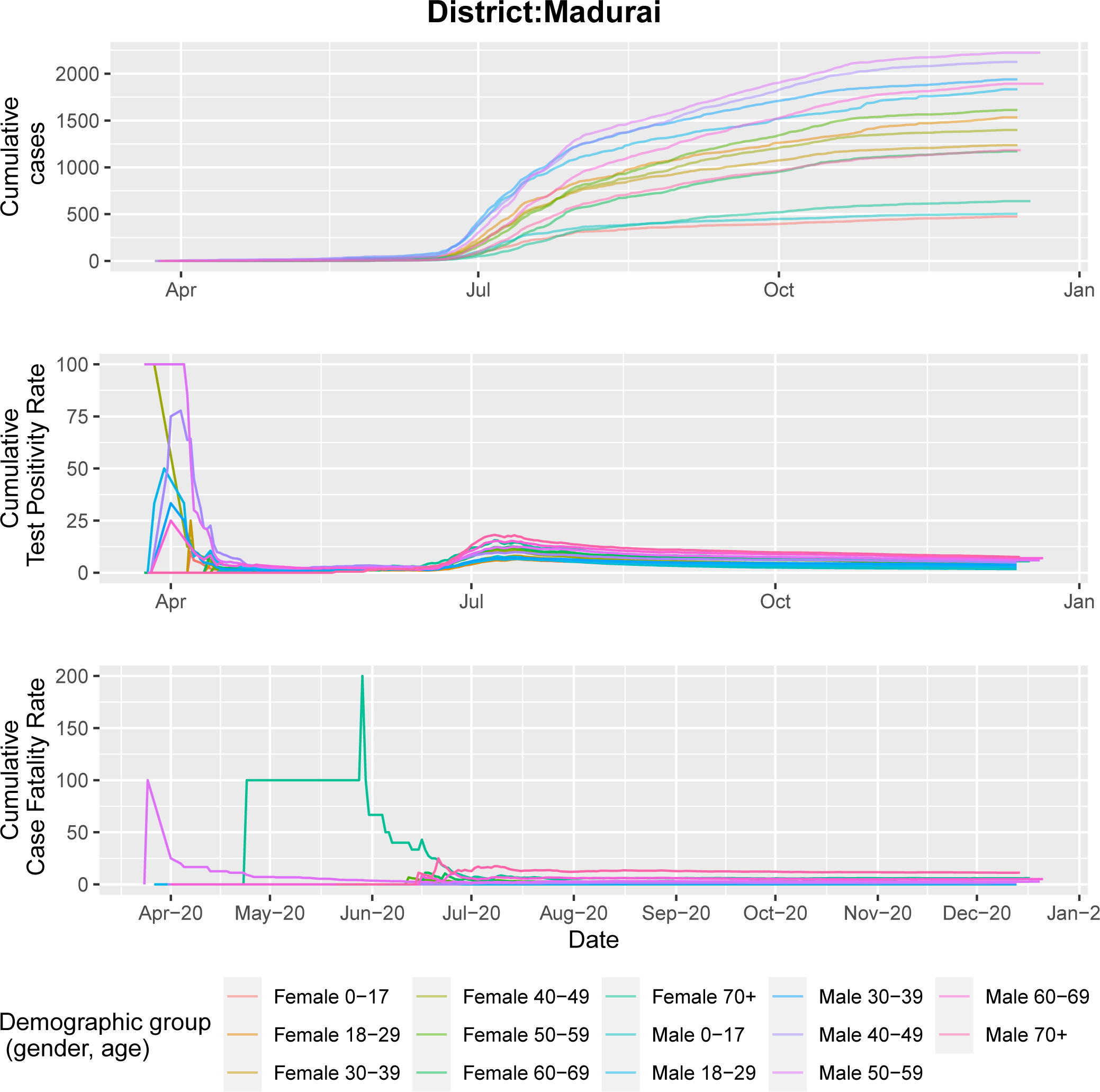
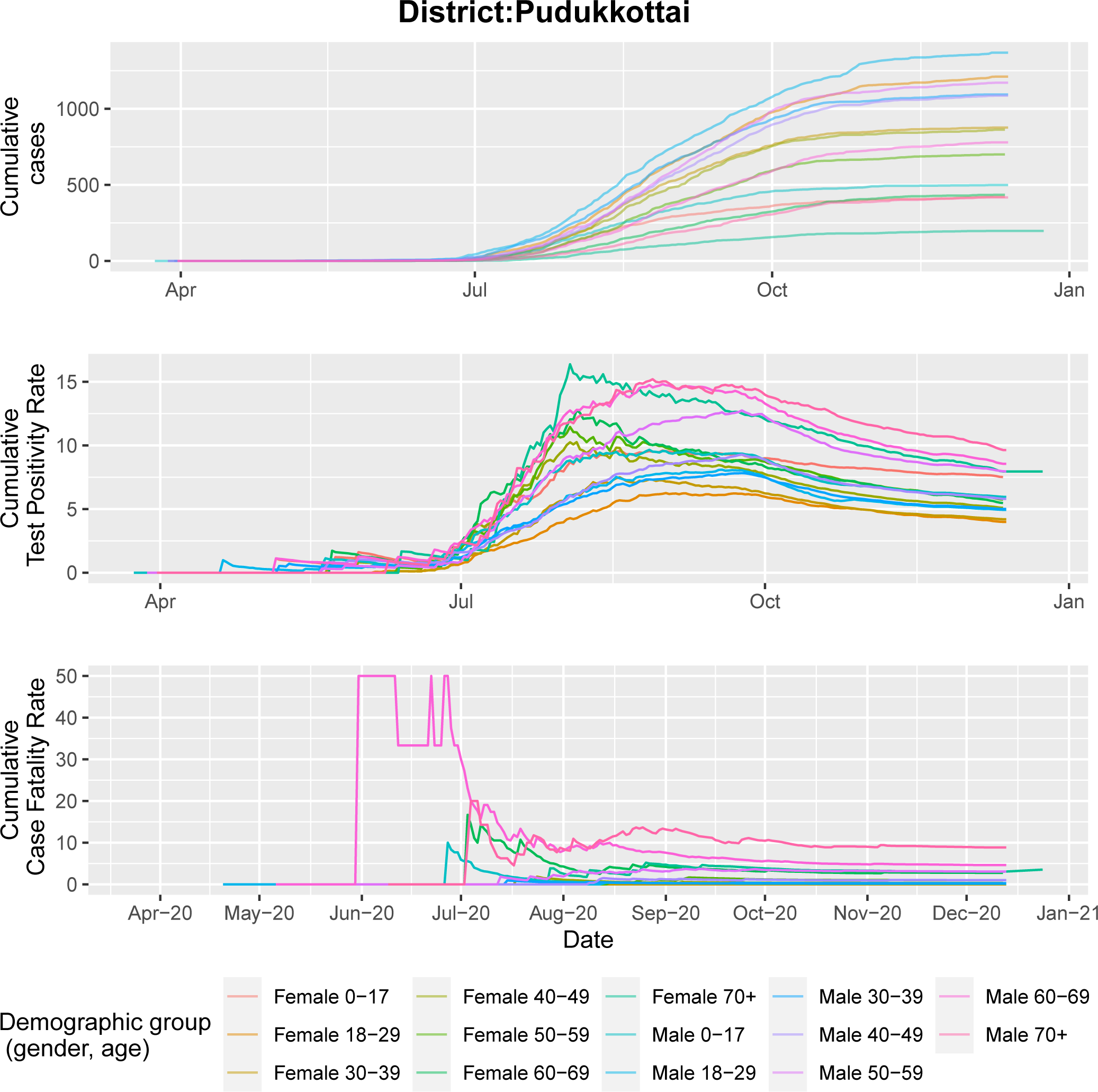
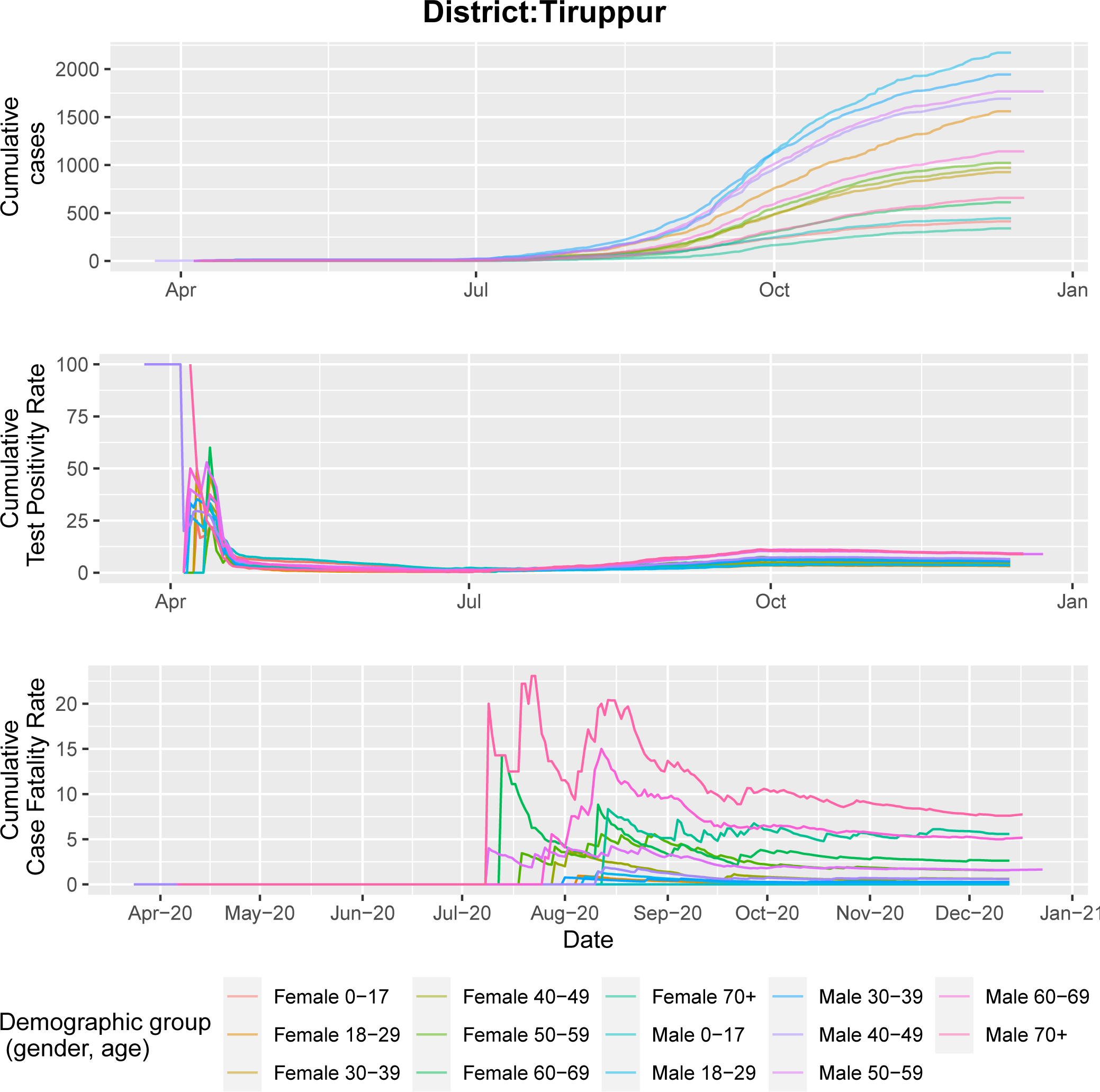
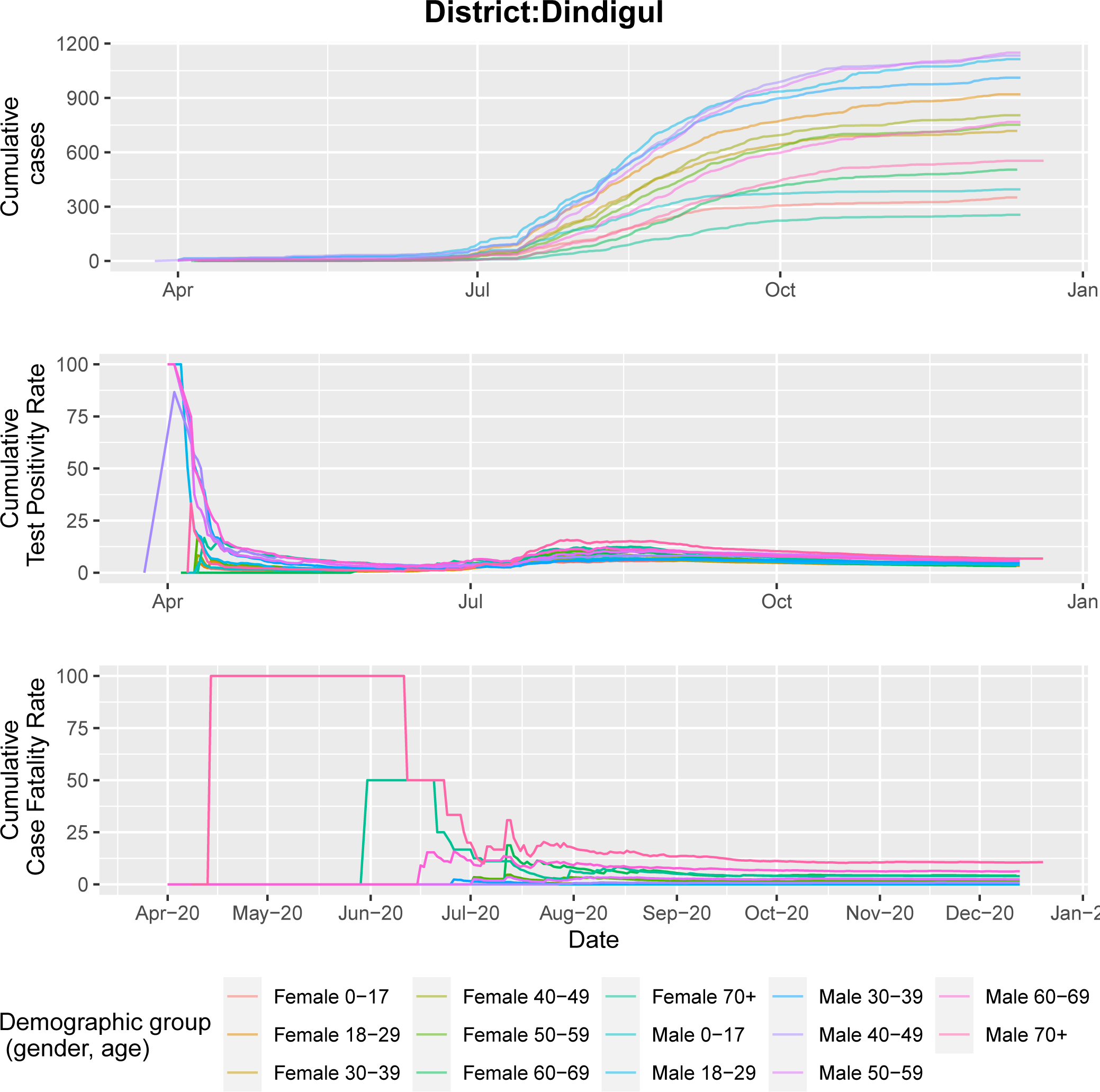
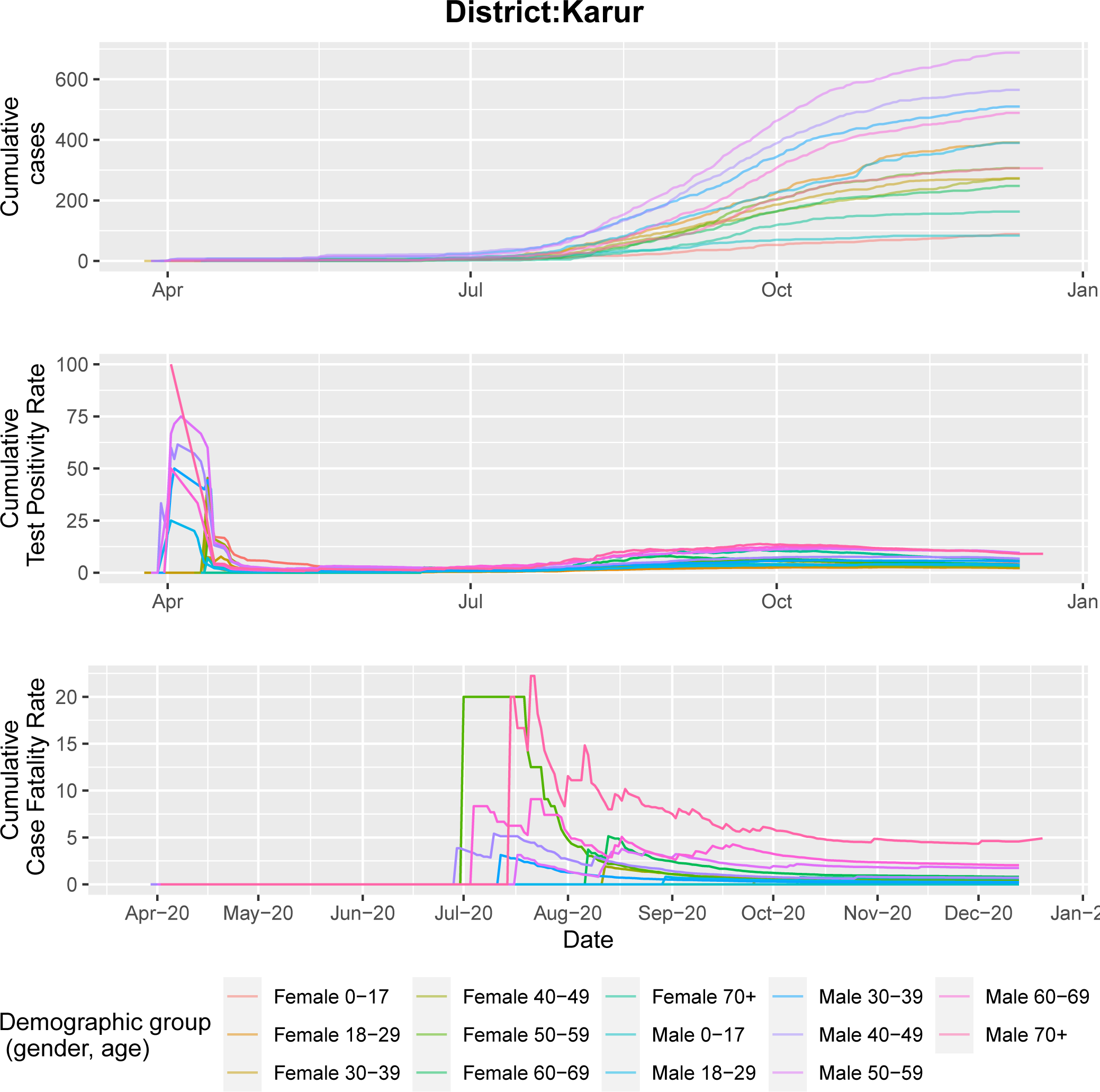
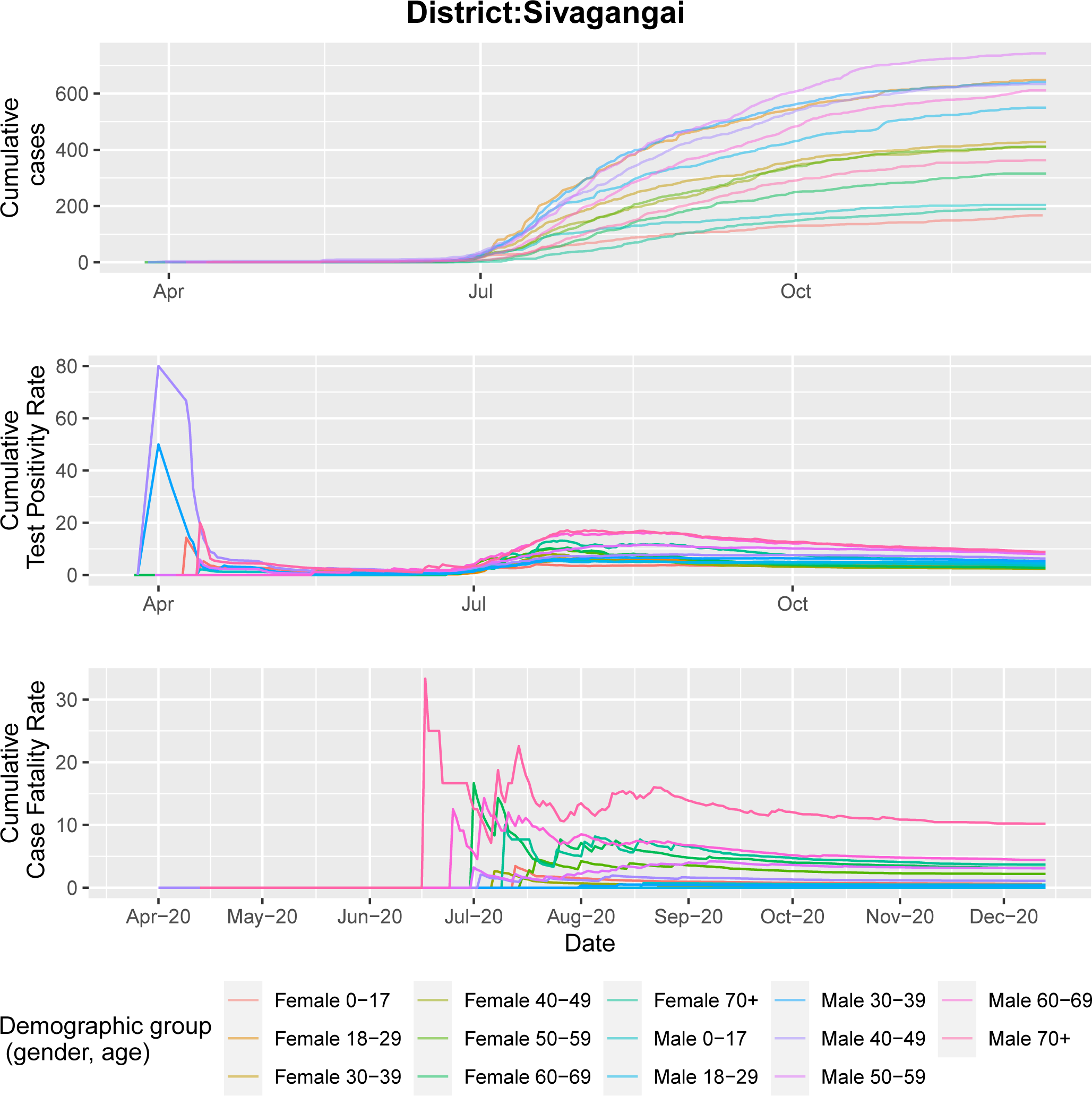
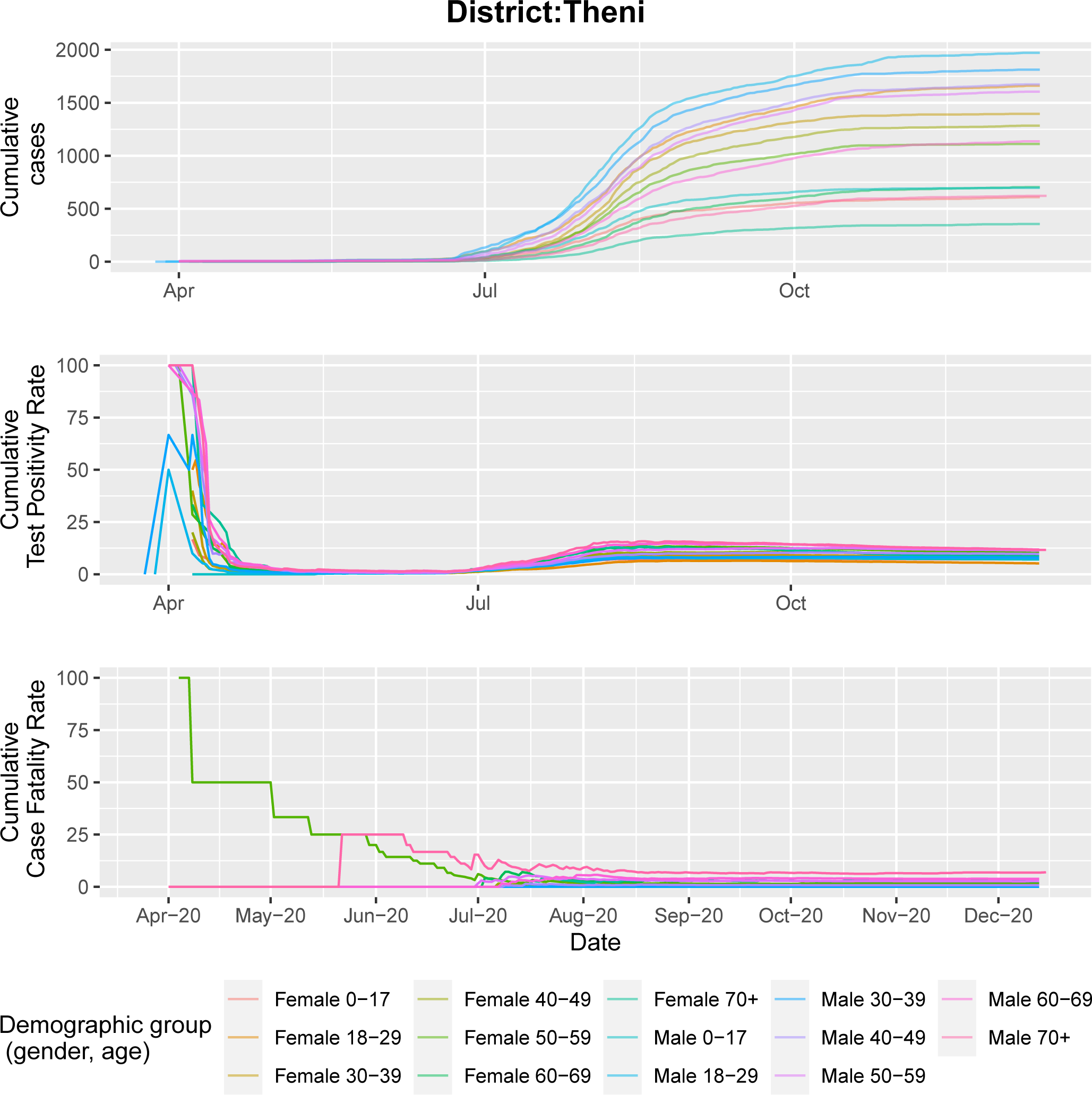
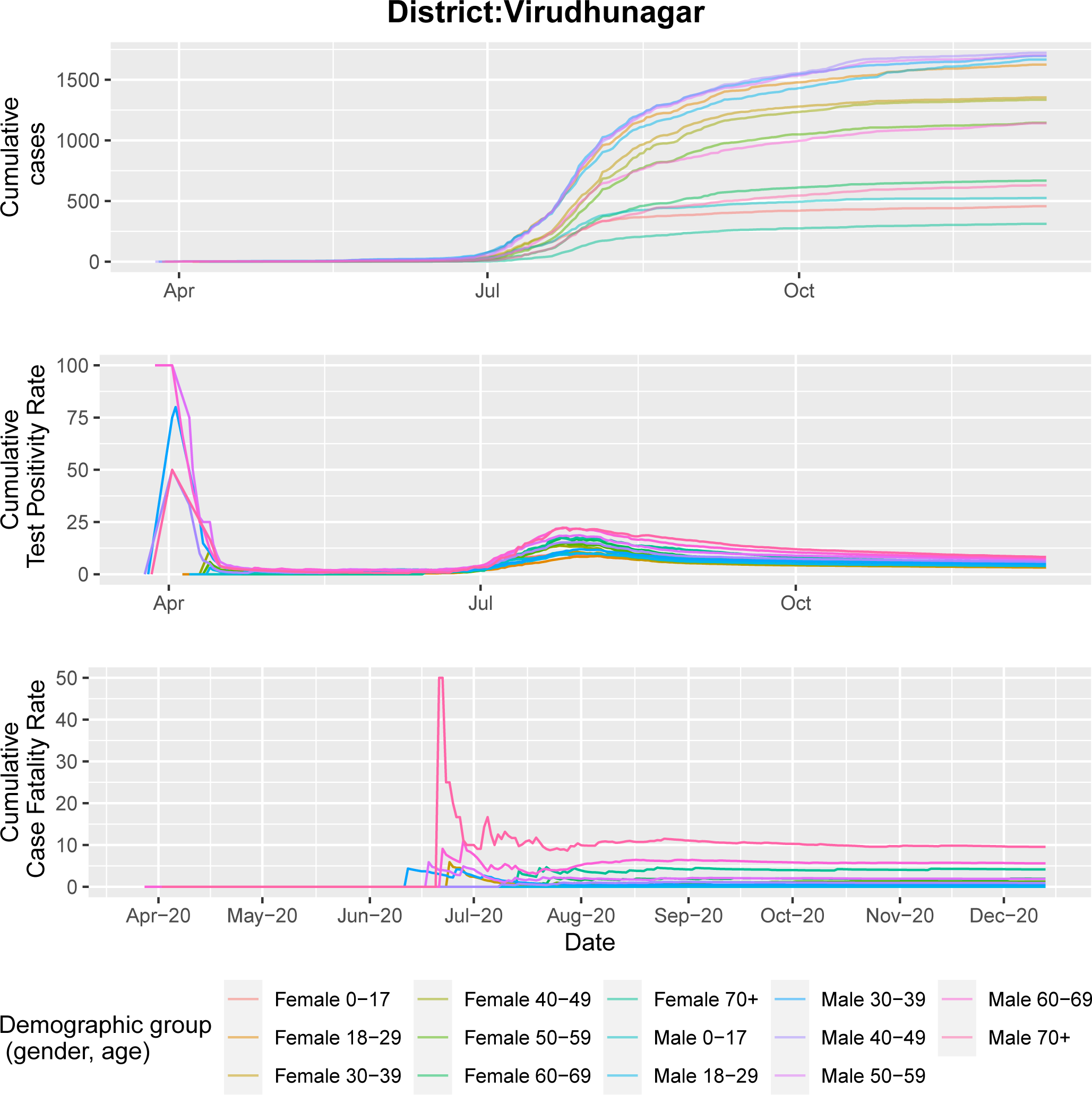
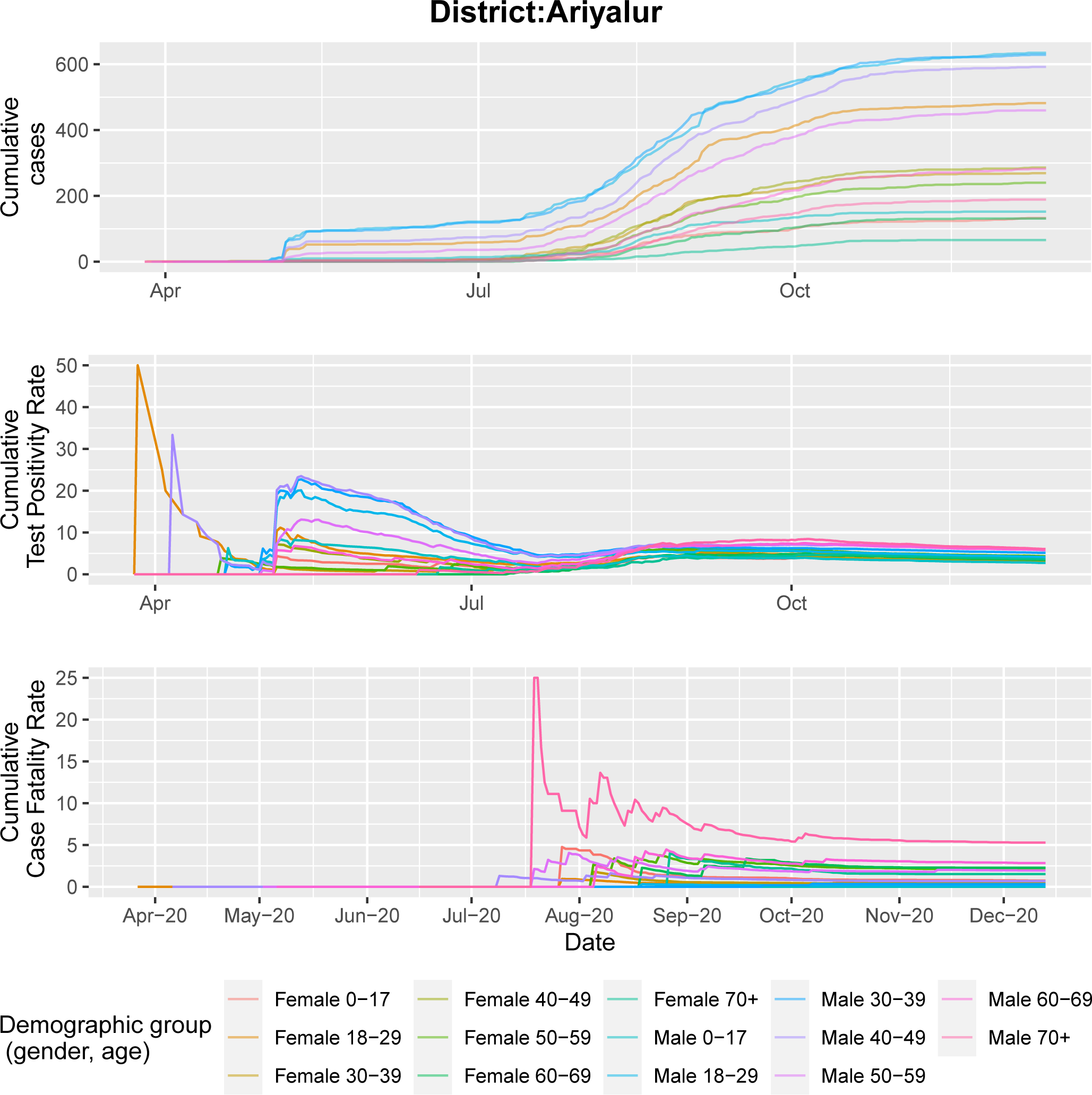
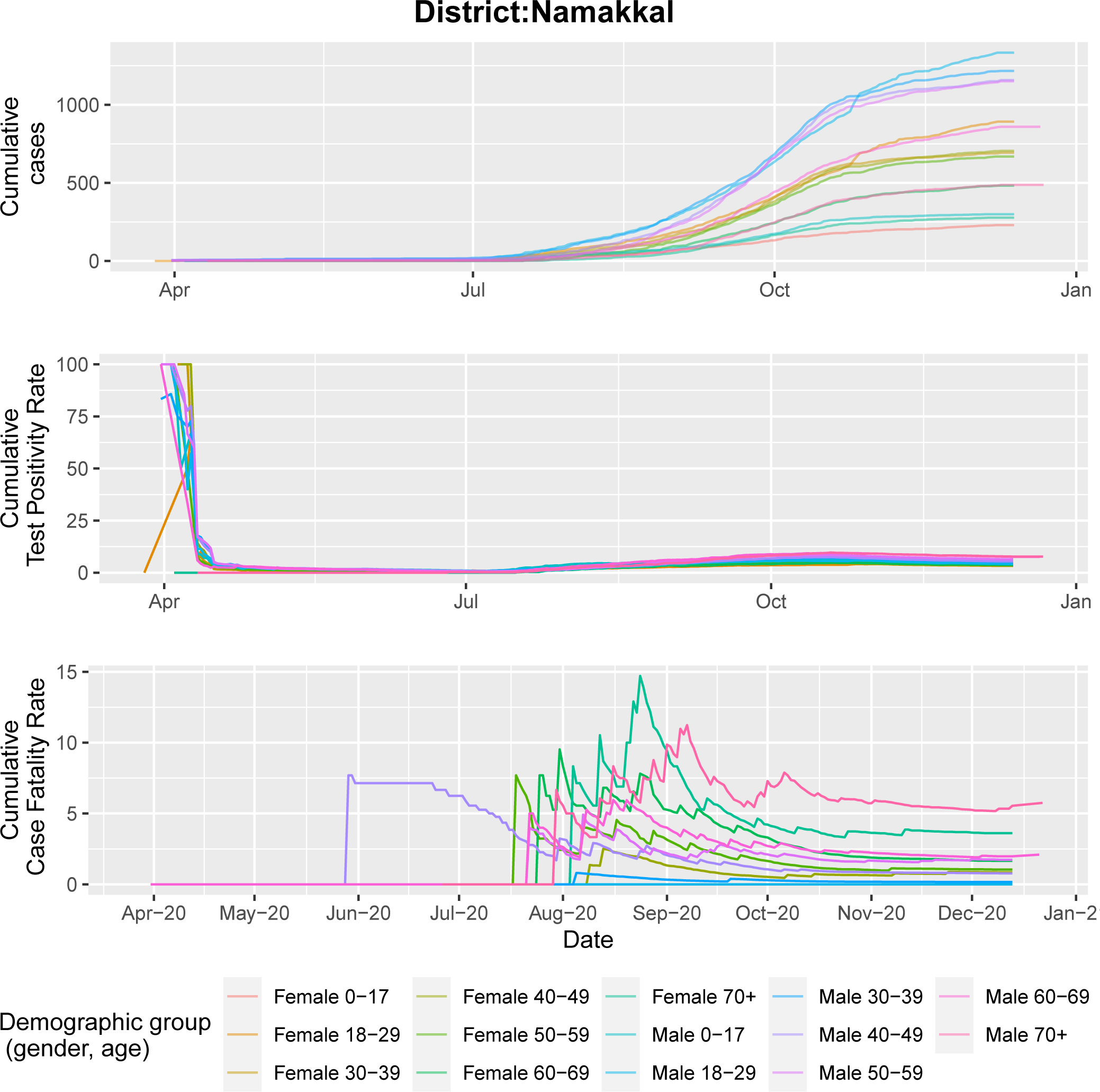
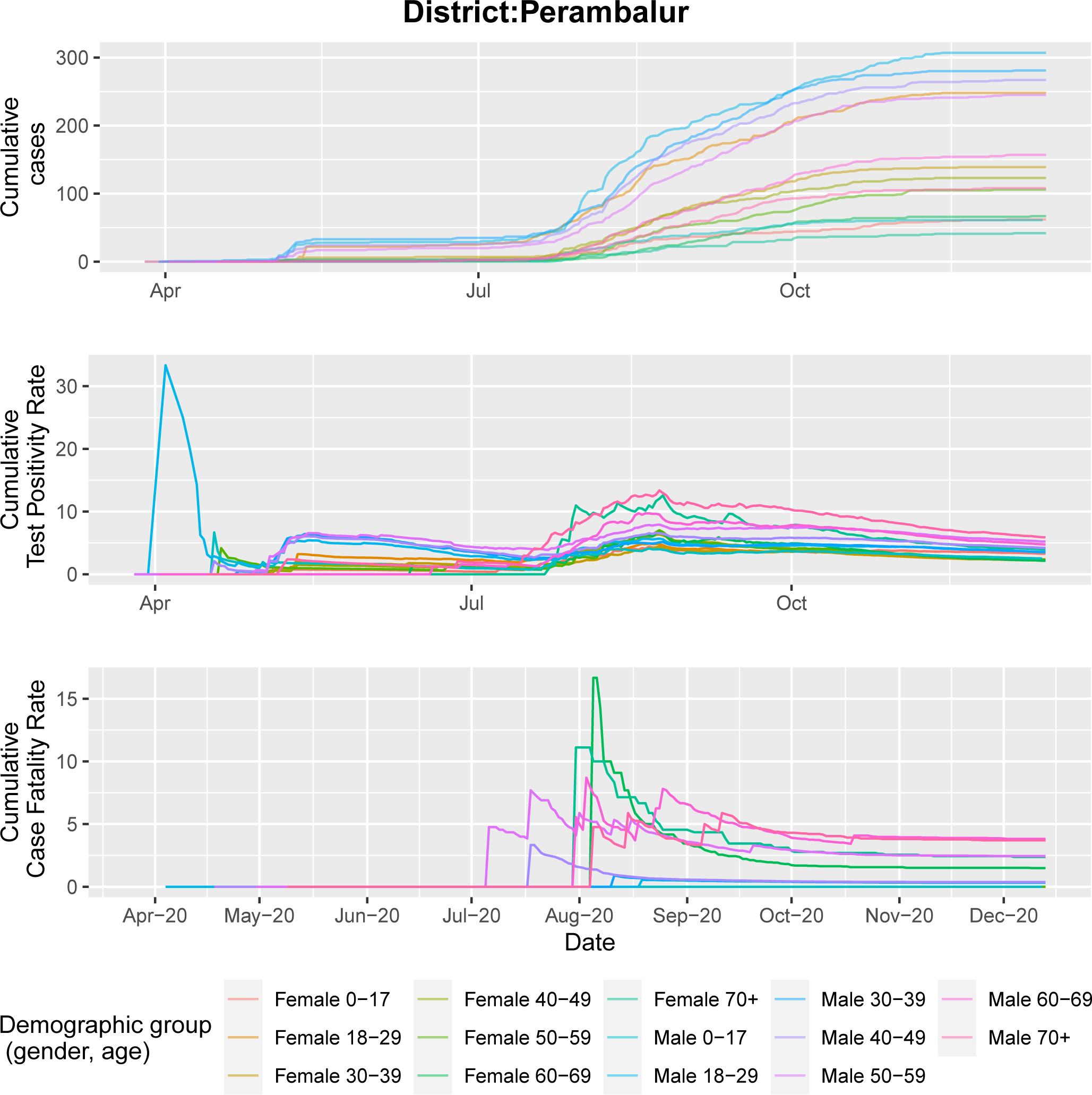
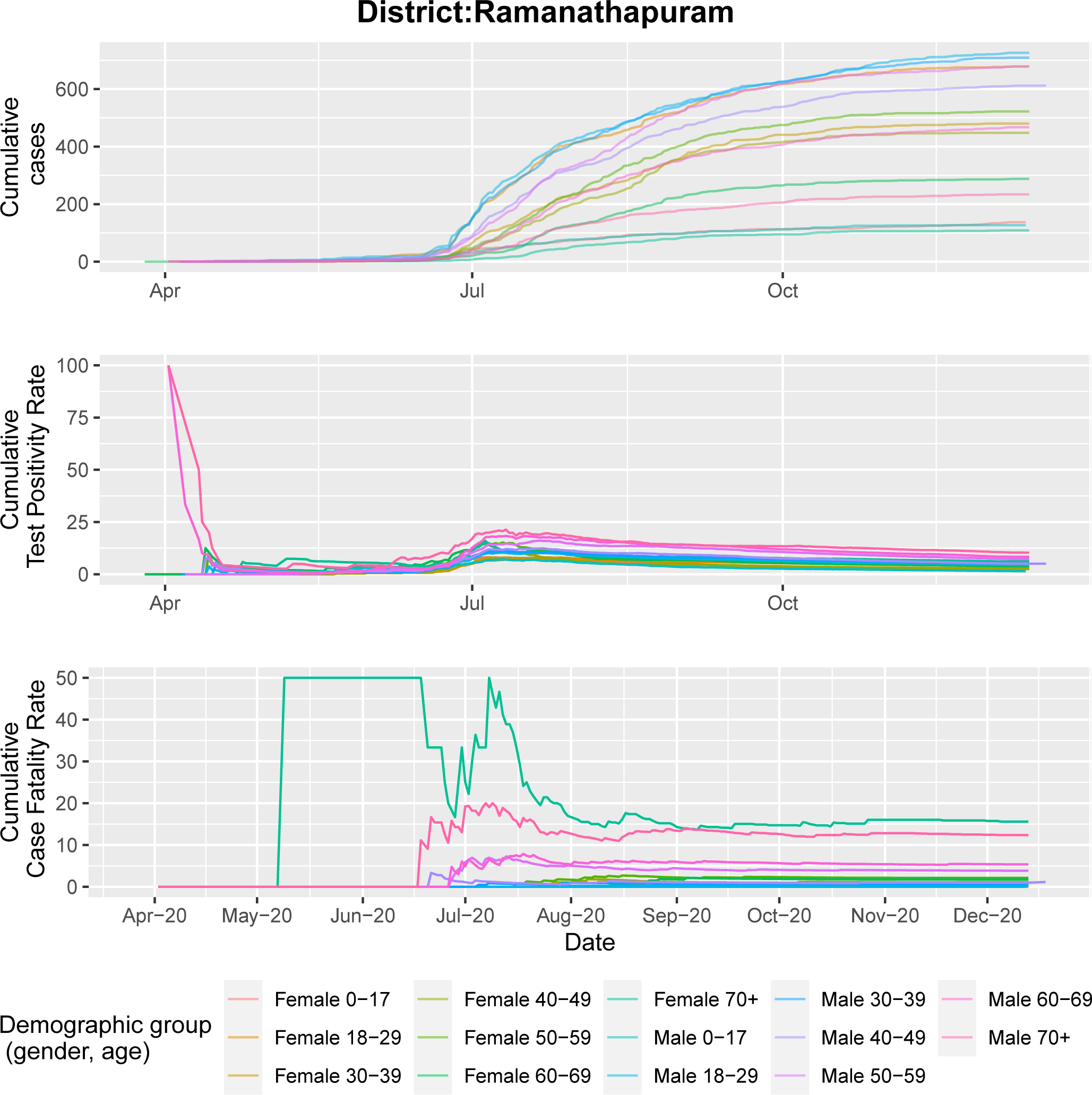
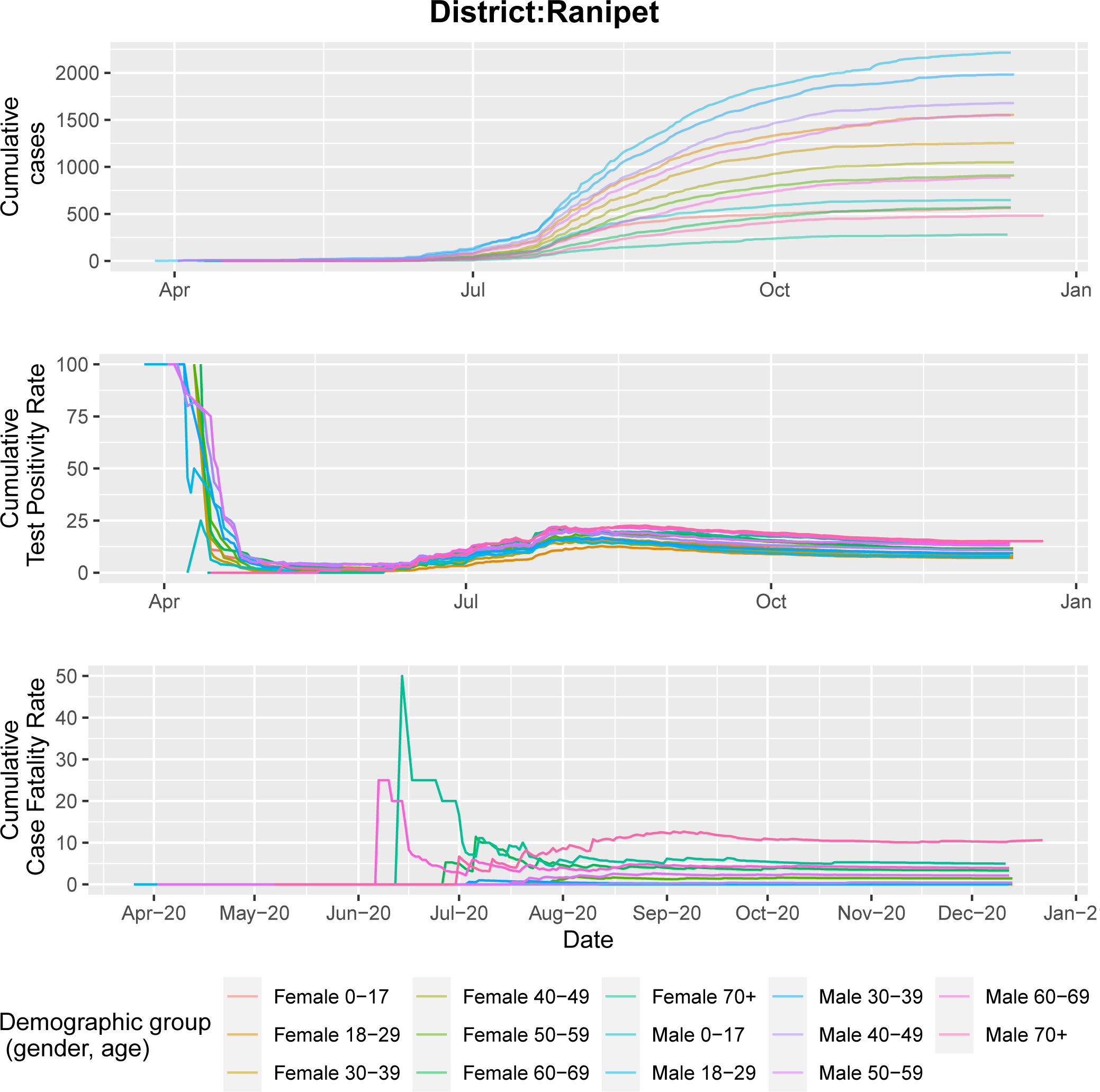
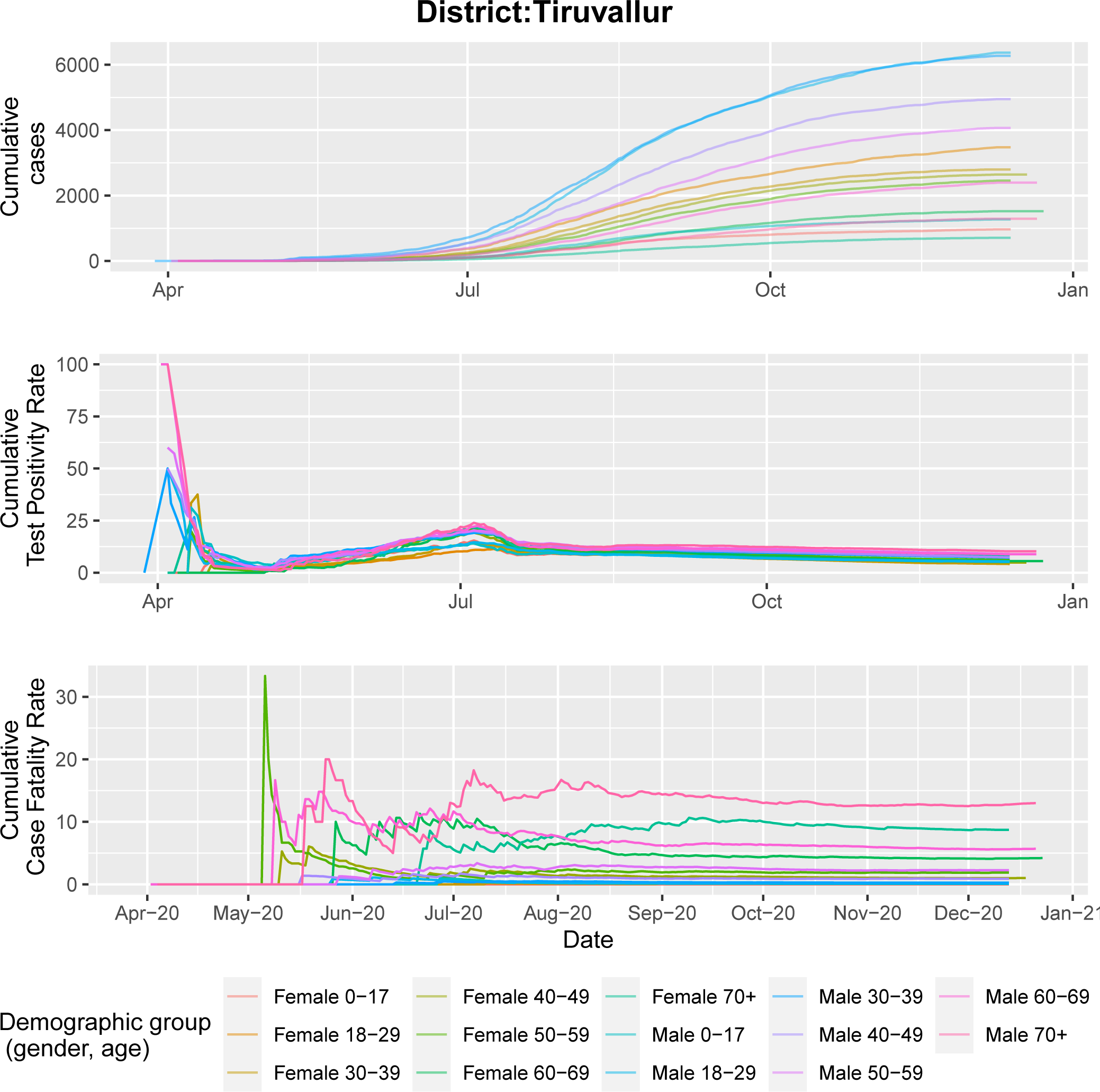
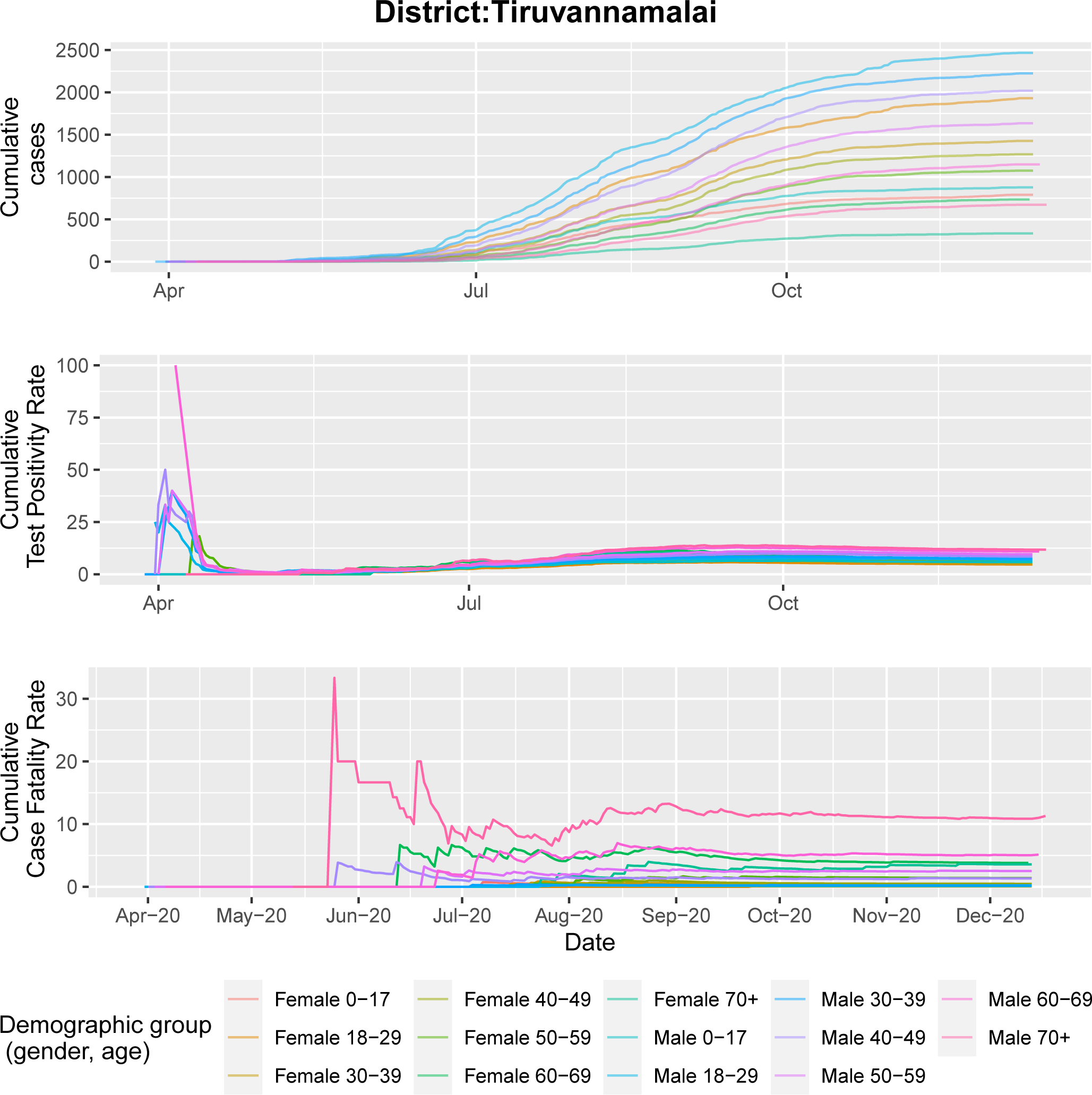
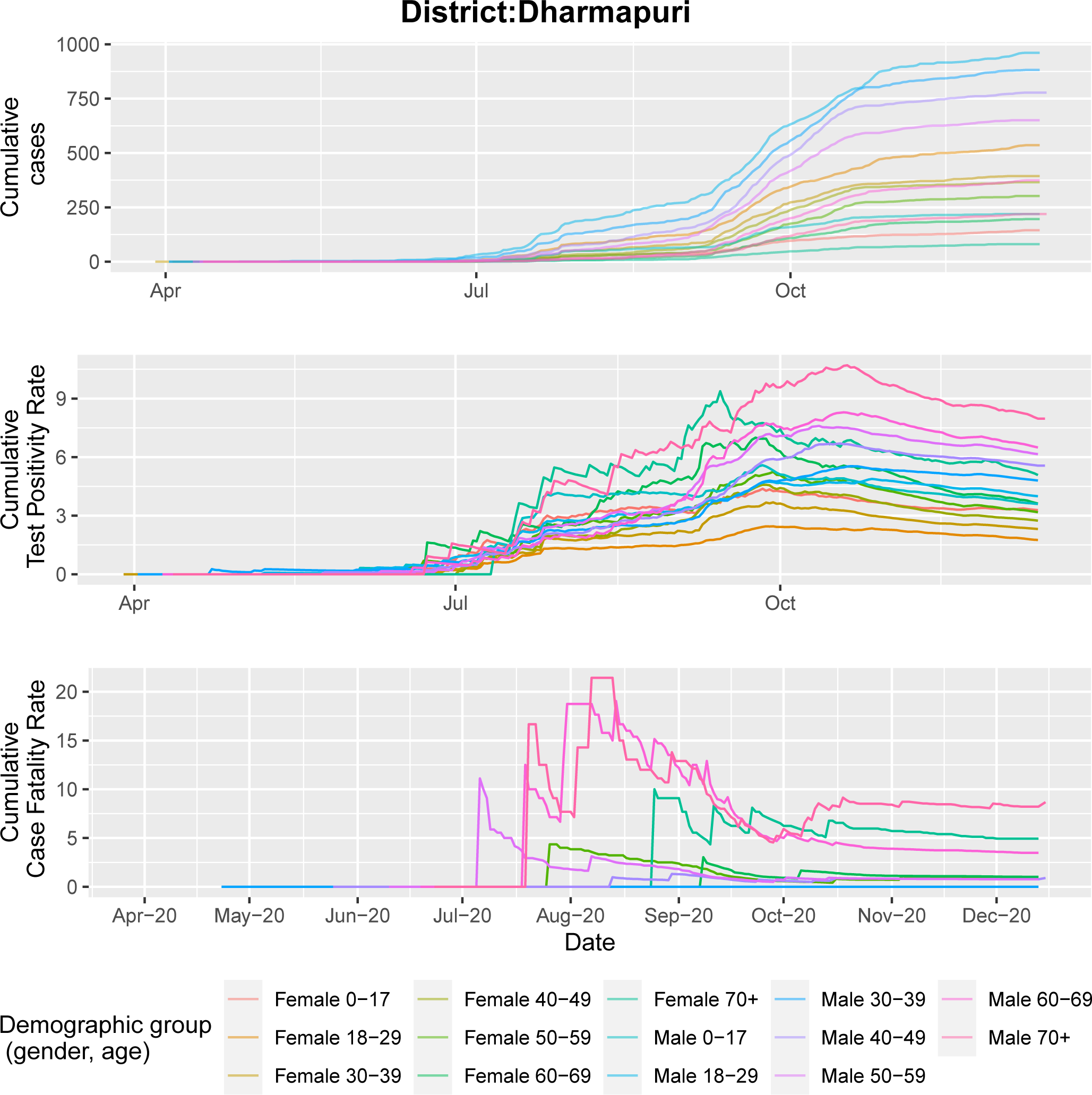
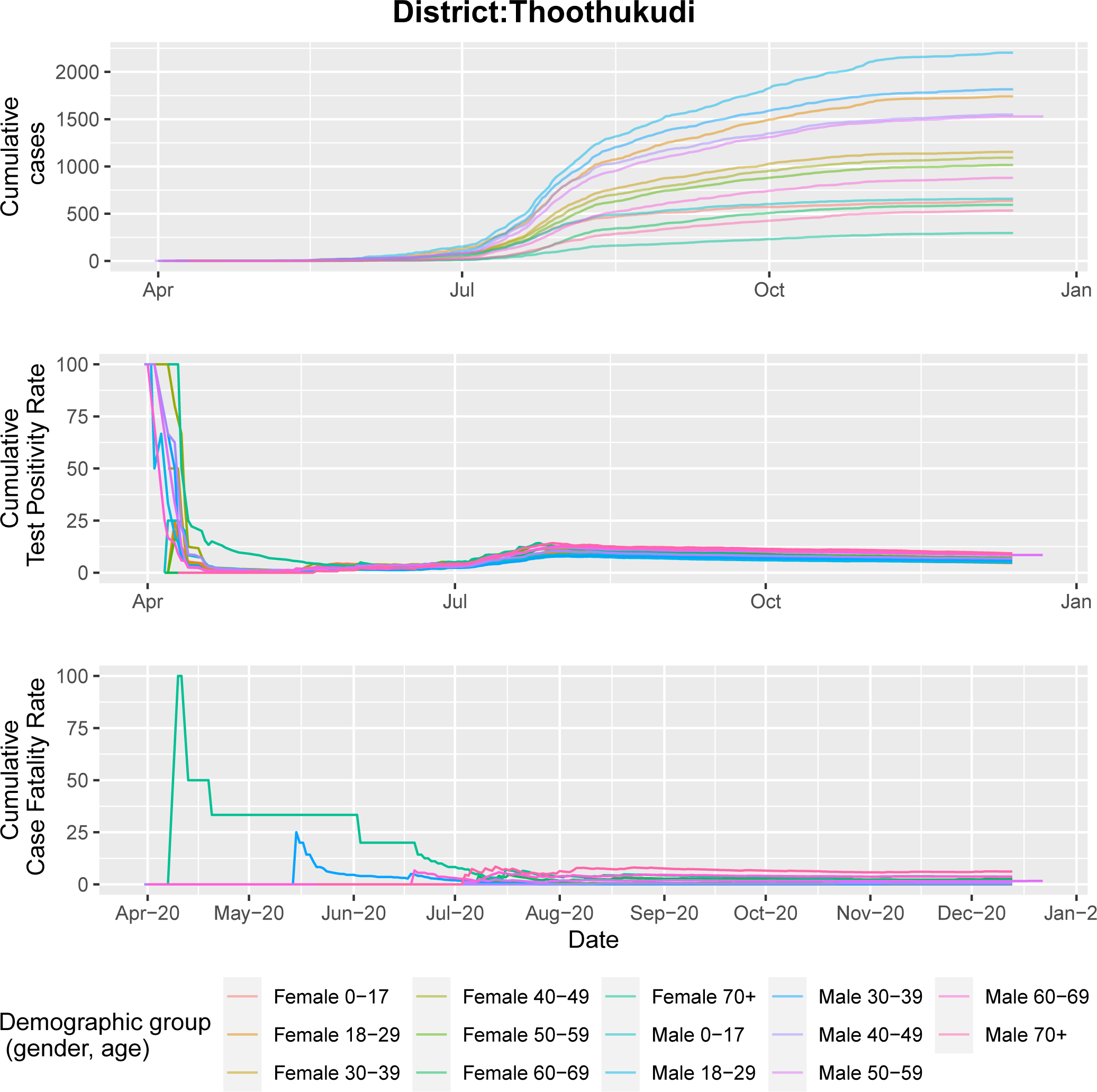
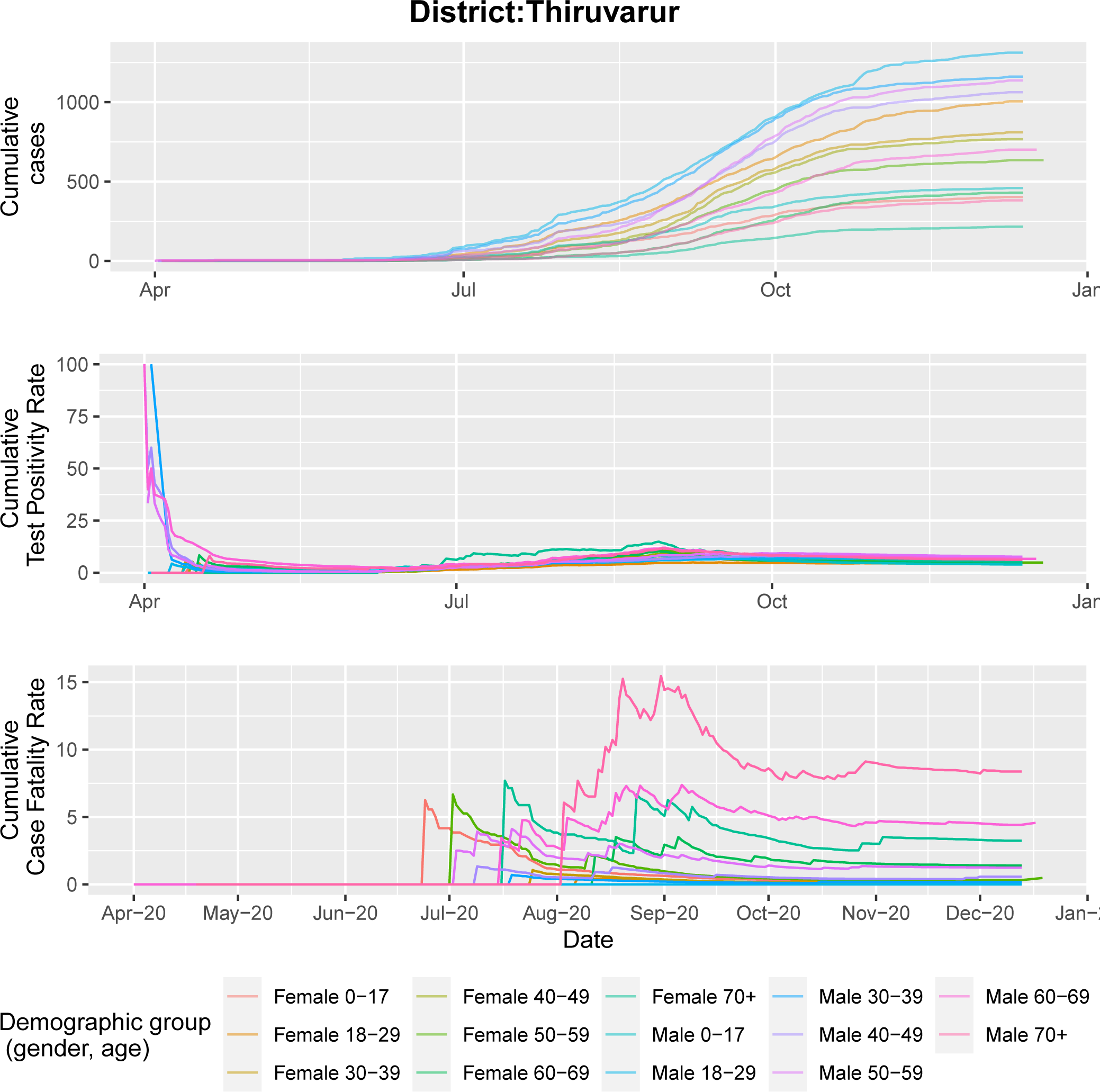
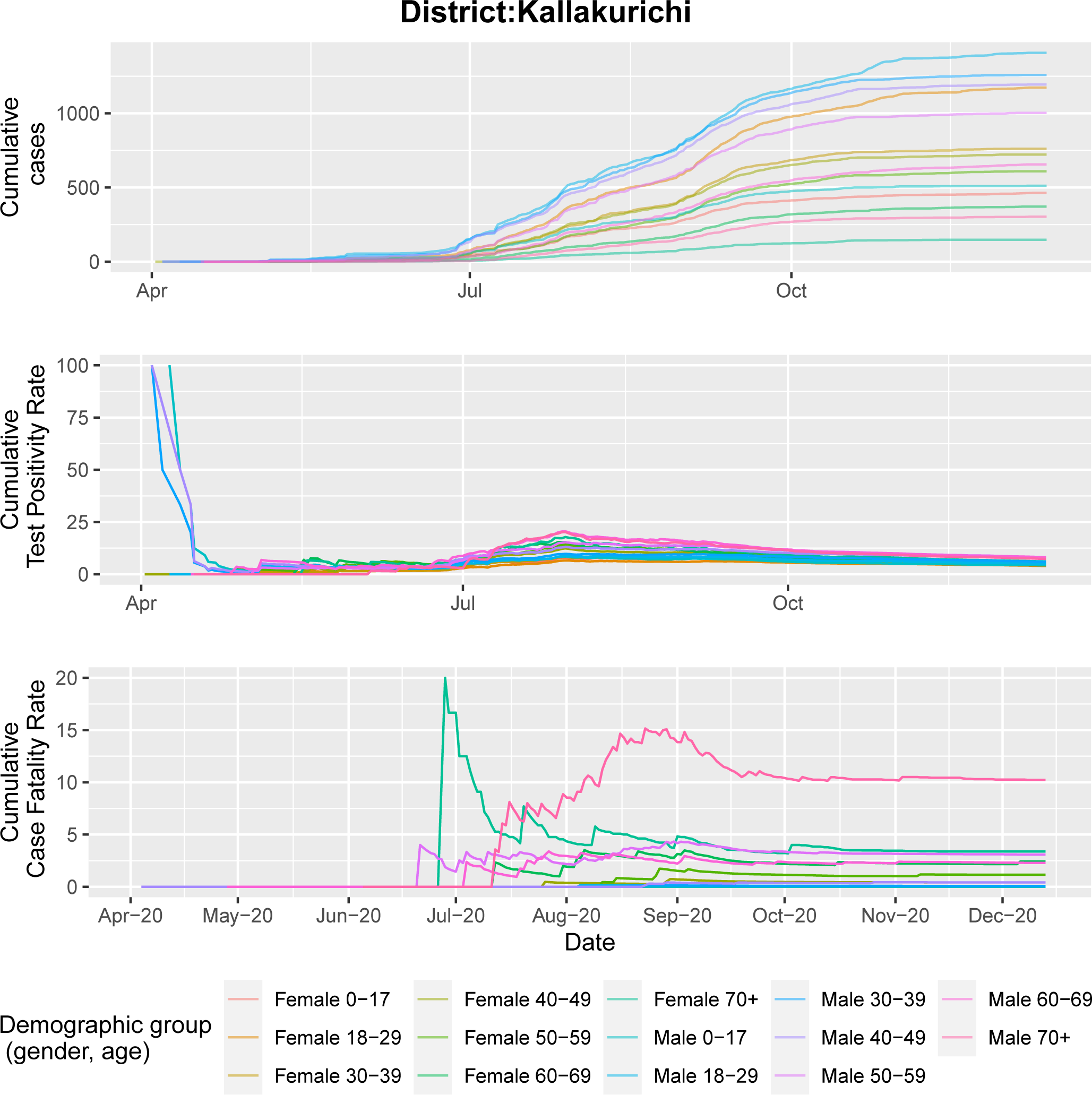
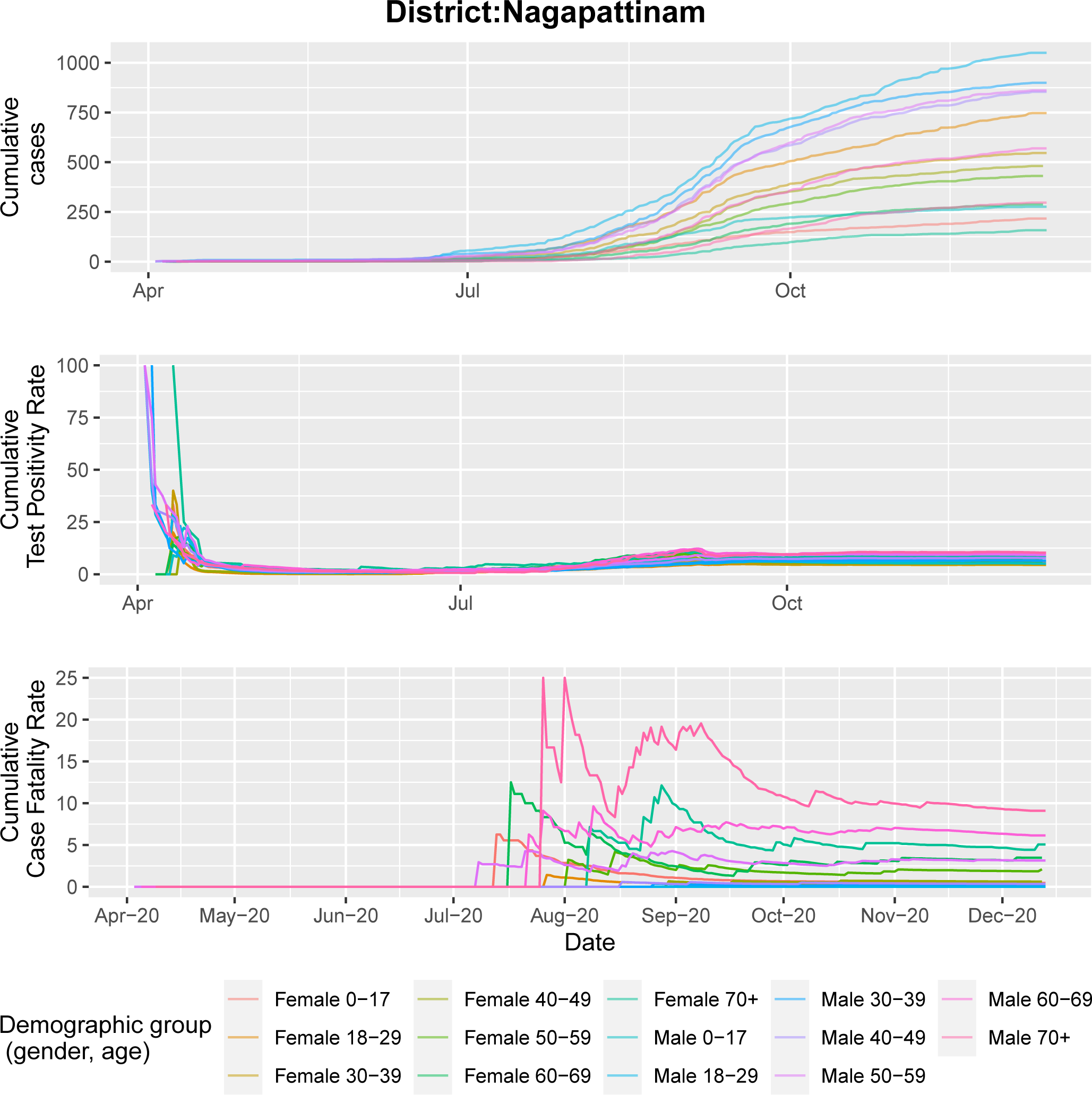
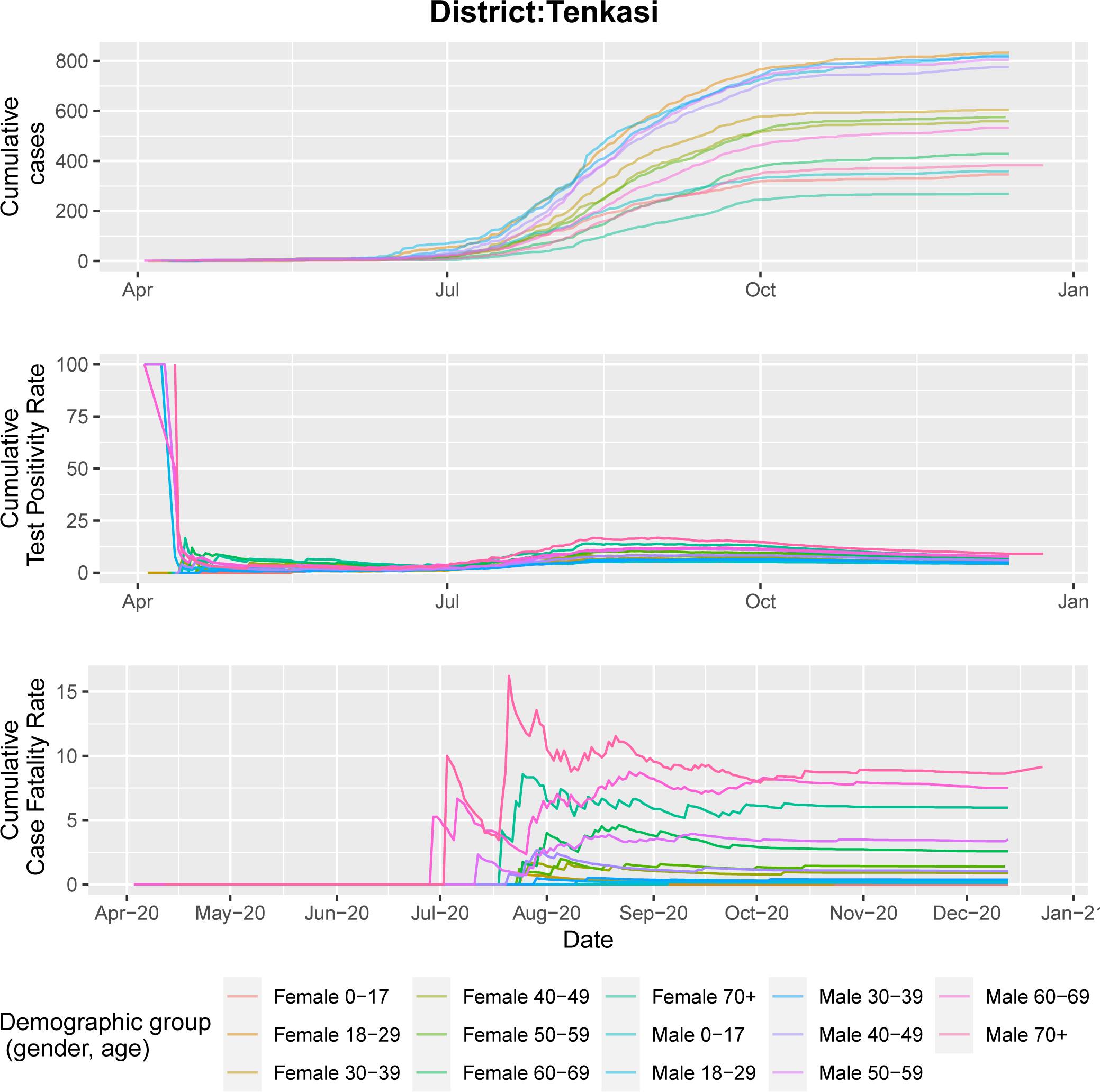
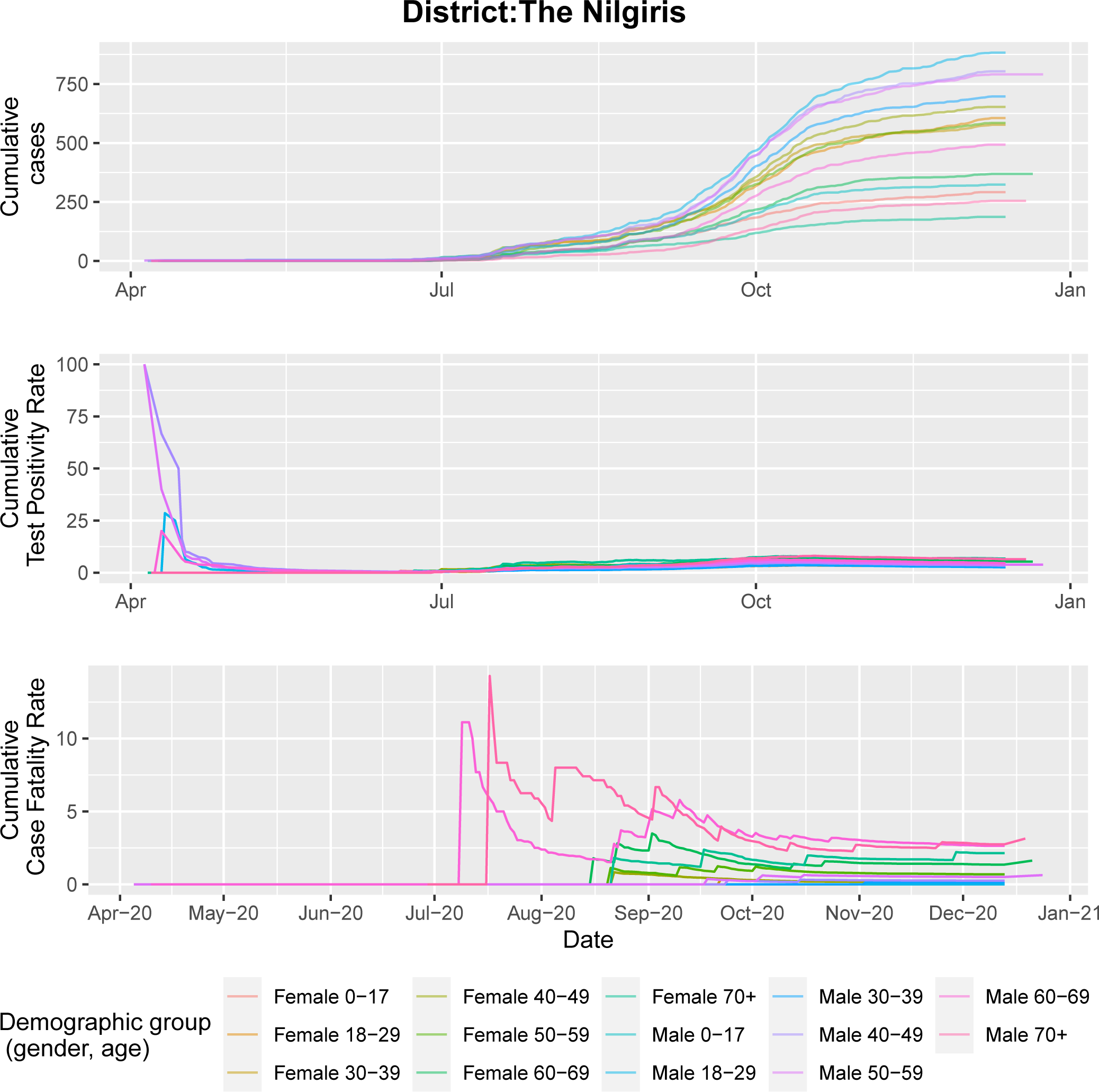
Cases, test positivity rate, and case fatality rate by demographic group and district.

## Data Availability

Aggregated data are reported in the Supplement.

